# Multimodal population-scale cell atlases define archetypes and extremes of human blood

**DOI:** 10.64898/2026.11.18.26368338

**Authors:** Olli Dufva, Shuang Li, J. Patrick Pett, Arianna Landini, Daniela Zanotti, Lisa Dratva, Lorenz Kretschmer, Krzysztof Polanski, Chuan Xu, Ioannis Sarropoulos, Ana-Maria Cujba, Jan P. Engelmann, Alessandro Raveane, Edoardo Giacopuzzi, Stephen Watt, Matiss Ozols, Giuditta Clerici, Manuel Tardaguila, Vasiliki Symeonidou, Paola Benaglio, Lucas Arnoldt, Beatrice Costa, Bruno Ariano, Sodbo Sharapov, Danai Vagiaki, Davide Bolognini, Eva Holtkamp, Andrei Cristea, Eljas Roellin, Dominique Von Schiller, Eve Coomber, May Hu, Gabriella Ivanova, Fereshteh Torabi, Nima Subasinghe, Laura Durrant, Chiara Chiereghin, Michela Colombo, Ilaria Gori, Cecilia Dominguez Conde, Matilde I. Conte, Blagoje Soskic, Alice Giustacchini, Ruth Eberhardt, Andrea Senacheribbe, Genes & Health Research Team, Nicole Staudt, John Wright, Richard Trembath, Craig A. Glastonbury, Julien Gagneur, Sarah Finer, Brian Clarke, Fabian J. Theis, Francesco Paolo Casale, Marc Jan Bonder, Fabiola Curion, George Vassiliou, Vivek Iyer, Oliver Stegle, Nicola Pirastu, David A. van Heel, Nicole Soranzo, Sarah A. Teichmann

## Abstract

The cellular composition of blood is fundamental to human physiology and is routinely measured to assess health and disease. Circulating immune cells show extensive heterogeneity, but the organising principles and extremes of variation across individuals remain undefined. Here, using single-cell transcriptomics across two biobank cohorts encompassing ∼6,000 individuals, we show that human blood occupies recurrent archetypal states defined by coordinated cell composition patterns. We delineate three major archetypes spanning naive lymphoid, myeloid and cytotoxic composition, and two minor archetypes characterised by type I interferon-driven inflammation and expanded premalignant CD5+ B cells. We develop cell2clone to infer lymphocyte clonal expansions, revealing how reactive clonal processes drive the cytotoxic archetype and premalignant clonal expansion establishes the CD5+ B cell archetype, creating outlier states that uncover context-specific effects of germline variants. Clonal haematopoiesis and mosaic chromosomal alterations influence blood states in an archetype-dependent manner. The archetype framework links blood states to common disease risk and clinical blood counts and, through a machine-learning model ArcheTypist, classifies infection and cancer immunotherapy outcomes. This population-scale framework has implications for diagnostics, early disease detection, immunotherapy stratification and mechanistic insights into disease susceptibility and therapy response, providing an organising model for identifying compositional subtypes across human tissues.

## Introduction

Human blood composition varies substantially across individuals, reflecting differences in immune history, ageing, environmental exposure, infection and somatic evolution. Single-cell transcriptomic studies have revealed extensive heterogeneity in circulating immune cells, identifying previously unappreciated cell populations and dynamic cell states associated with health and disease^1–5^. However, despite increasingly detailed catalogues of blood cell types, the organising principles underlying inter-individual blood variation are not well understood. Specifically, it is unknown whether variation in cell type abundance reflects largely independent fluctuations across cell types or if blood composition follows recurrent patterns of coordinated cellular variation. A unified framework of clinically meaningful blood subtypes could refine the concept of normal blood and reference ranges, and would allow placing individuals on trajectories of blood composition between health and disease.

Rare or expanded cell states have been identified in specific physiological and pathological contexts, including cytotoxic CD4⁺ T cells in supercentenarians^6^ and interferon-responsive myeloid states during viral infection^7^. However, it is not known whether such states represent isolated anomalies or instead occupy extremes of broader trajectories of blood composition across individuals. Population-scale atlases provide an opportunity to define normal tissue composition based on variation across the population, classify individuals based on cell composition and define outliers at the fringes of normal tissue composition.

Inter-individual variation, blood composition subtypes and extreme blood states could be driven by clonal expansion of cell types. Adaptive immune responses induce antigen-driven proliferation of lymphocytes^8^, while somatic alterations generate premalignant clonal expansions through myeloid^9^ or lymphoid^10^ clonal haematopoiesis and mosaic chromosomal alterations^11^. How these adaptive or premalignant clonal processes influence blood composition at population scale remains poorly understood. In addition to somatic alterations, rare outlier blood states generated by somatic clonal expansion may also create distinct regulatory contexts in which germline variants influence gene expression, contributing to cell type expansion and blood composition.

Understanding the organisation of blood variation has important clinical implications. The complete blood count (CBC) is a cornerstone of clinical medicine, providing a rapid and cost-effective readout of haematologic and immunologic status^12^. Measurements such as white blood cell count and leukocyte differentials are routinely used to detect infection, inflammation, haematologic malignancy and bone marrow dysfunction. Despite its clinical utility, the CBC compresses complex biological processes into a small set of aggregate measurements, offering limited insight into the cellular and molecular states underlying blood variation across individuals. Moreover, patient-specific blood count setpoints define disease trajectories^13^, but the underlying regulatory mechanisms and cellular and molecular processes maintaining these states or leading to deviations from them (e.g. leukocytosis, lymphocytosis and monocytosis^14^), remain unclear. Integrating molecular measurements at cellular resolution with conventional haematologic traits holds the promise of resolving the cellular and molecular mechanisms underlying both normal and abnormal blood states, bridging molecular and clinical phenotypes. Identifying subtypes of blood composition could provide a new approach to classifying the immune status of an individual, unlocking tools for predicting outcomes of infections and immune interventions.

Here, we leverage blood single-cell genomics of thousands of individuals across the UK Biobank^15^ (UKB) and Genes & Health^16^ (G&H) cohorts integrated with concurrent CBC measurements to show that human blood occupies recurrent archetypal compositional states defined by coordinated variation across immune cell populations (Fig. 1a). We propose that blood samples across a population can be grouped into archetypes with similar cell composition patterns, capturing coordinated variation across molecular and clinical measurements. We delineate trajectories toward extreme blood phenotypes and clonal and somatic processes that shape outlier blood states, define transcriptional programmes associated with these extremes, and uncover context-specific germline regulatory effects emerging in clonally expanded outlier cell states. Finally, we demonstrate how the archetype framework links blood immune states to common disease risk and can be used to explain clinical blood count abnormalities, classify COVID-19 infection outcomes and predict cancer immunotherapy-related adverse events. To support application of these approaches, we present computational tools for classifying blood and tissue archetypes (ArcheTypist, made available at github.com/TeichLab/ArcheTypist) and for adaptive lymphocyte clonality inference (cell2clone, available at github.com/TeichLab/cell2clone).

**Figure 1.**
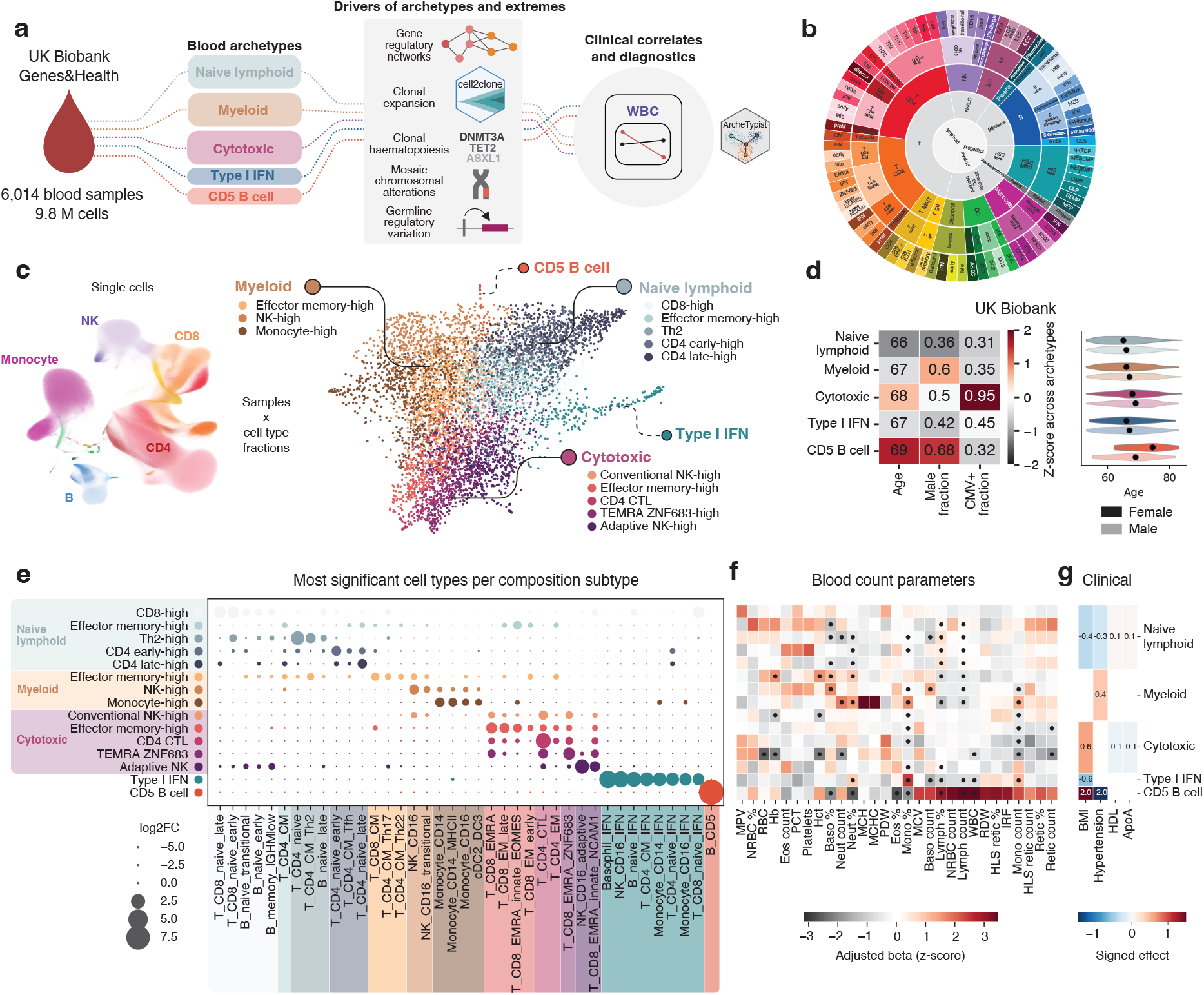
Blood composition archetypes across ∼6,000 individuals. **a,** Schematic of the study. **b,** Cell type annotation hierarchy, from inside to outside: lineage_1 (L1), lineage_2 (L2), celltype_1 (CT1), celltype_2 (CT2), celltype_3 (CT3). **c,** On the left, a UMAP representation of single cells in the CARDINAL scRNA-seq dataset, coloured according to CT3 annotations. On the right, a force-directed layout based on cell state fractions at the CT3 level across 6,014 samples in the UKB and G&H cohorts. Each dot indicates a blood sample from one donor. Dots are coloured by cell composition subtypes (Leiden clusters based on cell state fractions) annotated based on dominant cell types. **d,** Median age, fraction of male sex and fraction of predicted CMV seropositive donors in blood archetypes in the UKB cohort shown as heatmap. Distribution of age split by sex in blood archetypes is shown as a violin plot. **e,** Enrichment of cell types in the blood composition subtypes. Dot size indicates fold change relative to all other subtypes. **f,** Enrichment of complete blood count parameters in the blood composition subtypes after adjustment for age and sex in the UKB cohort shown as a heatmap. Significant associations are indicated with a dot. **g,** Enrichment of clinical risk factors in blood archetypes after adjustment for age and sex in the UKB cohort shown as a heatmap, with only significant associations shown.

In the future, blood archetypes based on our refined molecular and cellular analysis, potentially in conjunction with other clinical diagnostic modalities, can serve to better diagnose and identify early stages of disease. This paradigm also has potential for precision diagnostics of populations with distinct risk profiles for cardiometabolic, autoinflammatory and infectious diseases. Finally, we envision that our conceptual framework for blood presented here can serve as a prototype of identifying clinically relevant tissue composition subtypes beyond blood using emerging high-resolution datasets of increasing scale, for example population-scale spatial transcriptomics datasets of biopsies in tissues such as skin.

## Results

### Blood archetypes across human populations

To define the organising principles of inter-individual blood variation, we sought to identify subgroups across over 6,000 samples across 9.8 million peripheral blood mononuclear cell transcriptomes from the UKB (European ancestry) and G&H (British Pakistani and British Bangladeshi) cohorts (CARDINAL dataset, Giacopuzzi et al.^17^) based on the compositional space of co-varying cell types across 79 fine-grained celltype_3 (CT3) states (**Fig. 1b, Supplementary Fig. 1**). Graph-based clustering of samples based on cell state fractions identified 15 blood composition clusters or subtypes, which we annotated based on the most enriched cell types. Based on similarity of enriched cell types, we further grouped the composition subtypes into three major blood archetypes (**Fig. 1c-e, Supplementary Tables 1-2**): naive lymphoid (comprising composition subtypes enriched in naive CD8+, effector memory, Th2, and early and late naive CD4+ T cells), cytotoxic (including composition subtypes enriched in conventional NK, effector memory, cytotoxic CD4+, ZNF683^high^ terminally differentiated CD8+ T cells, and adaptive NK cells) and myeloid (including composition subtypes enriched in monocytes as well as effector memory T cell and NK cells). Additionally, two rare archetypes included one characterised by expanded CD5+ B cells consistent with monoclonal B cell lymphocytosis (MBL), a precursor state to chronic lymphocytic leukaemia^18,19^, and another defined by a type I IFN signature across multiple lineages, potentially implying inflammatory stimuli such as viral infection. We visualised the archetypes and composition subtypes on a force-directed graph, building a map of blood composition across populations (**Fig. 1c)**.

Blood archetypes showed distinct demographic associations. The naive lymphoid archetype was more predominant in younger individuals and females, whereas the myeloid archetype was more pronounced in males both in UKB and G&H, suggesting generalisability of the archetypes across populations with distinct ancestries and age structures (**Fig. 1d, Extended Data Fig. 1a, Supplementary Table 3**). Among the fine-grained composition subtypes, the naive lymphoid CD8+ T cell-high subtype showed the strongest association with young age in both sexes, suggesting a basal composition that can progress to other archetypes (**Extended Data Fig. 1a**). In contrast, the cytotoxic archetype increased with age and was strongly associated with cytomegalovirus (CMV) infection (**Fig. 1d**). Despite these associations, the archetypes showed substantial overlap in terms of age and sex, indicating that the archetypes capture variation beyond basic demographic factors. Our approach suggests that blood variation across populations can be broadly summarised between an axis defined by myeloid-biased and naive or cytotoxic lymphoid-biased archetypes, with more fine-grained composition subtypes identifiable in each of the archetypes.

### Archetypes are associated with blood counts and risk factors

To determine whether the archetypes correspond to clinically measurable features of blood, we examined their relationship with complete blood counts. The archetypes were reflected in complete blood counts, with the myeloid archetype showing higher neutrophil counts and the mature cytotoxic archetype showing higher lymphocyte counts (**Extended Data Fig. 1b,e**). These associations remained significant after adjusting for age and sex, indicating that the archetypes capture subgroups in blood beyond these factors (**Fig. 1f, Extended Data Fig. 1f-g**). The two rare archetypes were associated with more pronounced CBC alterations. The CD5+ B cell archetype showed markedly elevated lymphocyte counts, consistent with MBL. In contrast, the type I IFN archetype was associated with low lymphocyte counts and elevated monocytes, implying alterations induced by interferon signalling due to ongoing infection^20^.

We next asked whether blood archetypes could be driven by biased differentiation of haematopoietic stem cells (HSCs) favouring distinct lineages, supporting age-related myeloid bias^21,22^. Mature megakaryocyte–erythroid–basophil–eosinophil–mast progenitors (HSC_MPP_MEBEMP_L) were associated with the myeloid archetype, whereas the most immature HSC_MPP were associated with the naive lymphoid archetype (**Extended Data Fig. 2**). The type I IFN archetype showed depletion of mature MEBEMP progenitors, linking inflammatory blood states with circulating HSC states. Together, these findings imply that single-cell transcriptomics-derived archetypes can reveal the underpinnings of clinically observed blood counts, with links to HSC output.

To evaluate whether blood archetypes are linked with clinical phenotypes beyond blood cells, we correlated archetypes with risk factors and common diseases using biobank metadata and health records, adjusting for age and sex. In UKB, the cytotoxic archetype was associated with higher body mass index and lower high-density lipoprotein cholesterol and apolipoprotein A1 (**Fig. 1g, Extended Data Fig. 1f**). The myeloid archetype was associated with hypertension, whereas the naive lymphoid archetype showed opposite patterns to both the myeloid and cytotoxic archetypes. In the younger G&H population, the myeloid archetype was similarly associated with hypertension, hypercholesterolemia and other diseases linked to adverse metabolic health such as cholelithiasis (**Extended Data Fig. 1c-d, Supplementary Tables 3-4**). Consistent with UKB, a subtype enriched with CD4+ CTLs (cytotoxic T lymphocytes) in the cytotoxic archetype was associated with hypertension in G&H. Associations of individual cell types and states with disease are explored in more detail in Zanotti et al.^23^ Together, these associations indicate a link between blood archetypes and metabolic health, with lower cardiovascular disease risk factors in individuals with the naive lymphoid blood archetype.

### Gene regulation underlying archetype trajectories

To chart the limits of blood compositional space, we sought to define trajectories that position individuals along the major axes of blood variation (**Fig. 2a**). We inferred trajectories and pseudotime across samples (instead of cells) using scFates^24^ with cell state fractions as the feature space, defining the CD8+ T cell-high naive lymphoid subtype as the trajectory root based on its enrichment in young individuals. From this starting point, trajectories progressed through a central intermediate region of the cell composition embedding before branching towards extremes in each archetype, with increasing dominance of specific immune lineages and states with increasing pseudotime.

**Figure 2.**
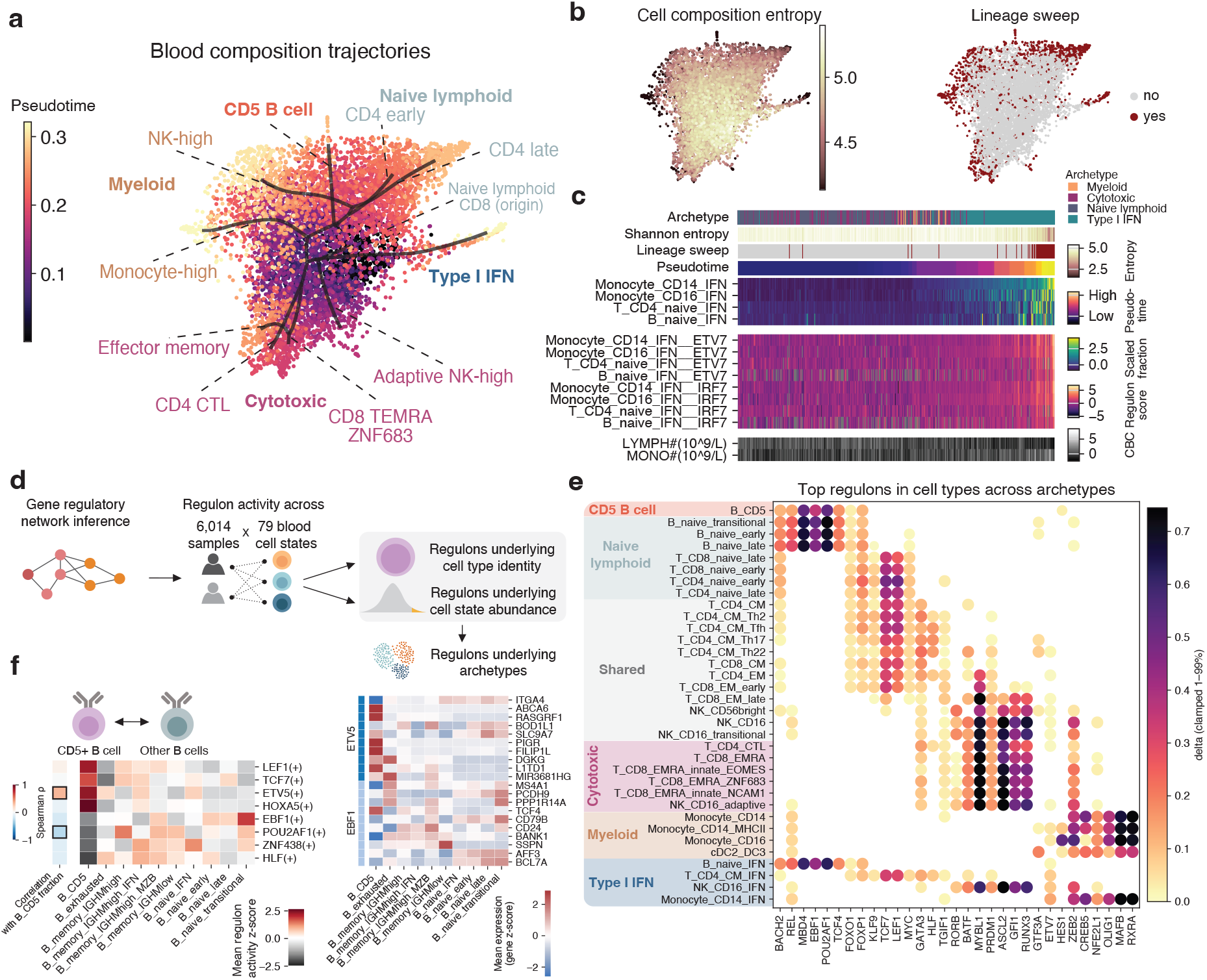
Blood composition trajectories and underlying gene regulatory networks. **a,** Cell composition trajectories shown on a force-directed layout of cell state composition coloured based on blood composition pseudotime. **b,** Shannon entropy of cell state fractions and lineage sweep status (defined as entropy < 4.5) for each sample coloured on the force-directed layout. **c,** Heatmap of features correlating with the type I IFN blood composition trajectory, including archetype, blood composition entropy, lineage sweep status, blood composition pseudotime, and selected highly correlating cell type fractions, regulons and CBC parameters. **d,** Schematic of the GRN inference workflow. **e,** Heatmap of cell type-specific regulons for selected archetype-enriched cell types at the CT3 level. Colour indicates the delta of mean scores of differential enrichment of a regulon in each cell type compared to all others. Cell types are grouped based on the archetype they are enriched in and regulons are hierarchically clustered. **f,** Heatmap of selected differentially active regulons between CD5+ B cells and other B cells, and of ETV5 and EBF1 regulon targets with highly variable expression between shown cell types. Column on the left indicates correlation of the regulon activity with CD5+ B cell fraction.

To robustly quantify the extent of skewing of the cell state repertoire towards few or single cell states, we computed Shannon entropy of sample-level cell type composition (**Fig. 2b**), where high entropy corresponds to ‘normal’ blood where cell types are evenly distributed. Entropy decreased progressively along the blood composition trajectories, correlating inversely with pseudotime and defining a subset of individuals with an outlier low entropy, with few cell states dominating their blood composition. We termed this loss of blood diversity ‘lineage sweep’, reflecting the dominance of one lineage (**Fig. 2b**). Although entropy decreased with age, age alone did not explain the most extreme decreases in entropy (**Extended Data Fig. 3a**). Entropy was markedly decreased in the CD5+ B cell archetype, where terminal trajectory states included individuals in whom CD5+ B cells comprised more than half of blood mononuclear cells (**Extended Data Fig. 3b**). Additionally, several other subtypes included a minor subset of low-entropy donors, indicating that all archetypes can progress to cell composition extremes where one or few cell types dominate in blood, as exemplified along a trajectory towards the type I IFN archetype with decreasing CBC lymphocyte and increasing monocyte counts (**Fig. 2c**). Together, these findings indicate that blood archetypes are composed of continuous trajectories of coordinated immune variation towards extreme compositions that reflect changes in the CBC.

A key advantage of transcriptomics is the ability to understand underlying regulatory mechanisms that govern cell states, such as those constituting blood archetypes. We performed gene regulatory network (GRN) inference using SCENIC^25^ to understand the cell type-specific GRNs underlying the identity and abundance of the cell types defining archetypes (**Fig. 2d**). We inferred 347 regulons using a subsampled dataset and scored these regulons on sample-specific pseudobulks at the CT3 level, allowing us to identify regulons enriched in given cell types and correlate regulon activity with cell abundance and composition trajectories.

We first assessed regulons associated with cell type identity showing higher activity in a given cell type compared to all others (**Fig. 2e, Supplementary Fig. 2-3, Supplementary Tables 5-7**). Cell type-specific TF activities included MSC, FOXD1/2 and NFIA activity distinguishing adaptive NK cells from other NK subsets (**Extended Data Fig. 3d**) and the interferon-responsive TFs ETV7 and IRF7 enriched in type I IFN archetype-associated cell states across lineages (**Fig. 2c**). CD5+ B cells displayed increased activity of regulons including LEF1, TCF7, ETV5 and HOXA5 relative to other B cells and conversely reduced activity of canonical B lineage factors EBF1 and POU2AF1 controlling genes known to be downregulated in MBL, indicating regulatory rewiring that diverges from conventional B cell differentiation (**Fig. 2f**).

Next, we correlated regulon activity with fraction of the same cell type across samples to ask which regulons may underlie variation in cell abundance across the population and therefore the archetypes enriched with these cell types (**Extended Data Fig. 3c, Supplementary Table 8**). RXRA and MAFB activity was associated with CD14+ monocyte abundance, suggesting importance in establishing the myeloid archetype, consistent with these transcription factors (TFs) driving monocyte differentiation^26–28^. EOMES, RUNX3 and GATA3 activity correlated with increased CD4+ CTL abundance, suggesting relevance for the cytotoxic archetype (**Extended Data Fig. 3e**). Increased ETV5 activity correlated with the CD5+ B cell fraction, implying that ETV5 may both divert CD5+ B cell identity from other B cells and promote increasing CD5+ B cell counts, consistent with progression from low-count to high-count lymphocytosis (**Fig. 2f, Extended Data Fig. 3c**). Together, these findings provide insights into regulatory mechanisms controlling blood archetypes and mechanisms of premalignant progression, with potential implications for diagnostics and early interventions.

### Clonal expansions shape blood archetypes

Progression toward archetypal blood extremes was accompanied by expansion of specific cell states, which may result from clonal proliferation due to antigen recognition by adaptive lymphocytes or somatic alterations conferring a growth advantage. We next sought to evaluate the prevalence, cellular distribution and phenotypic consequences of clonal expansion across individuals and blood archetypes.

We developed cell2clone, a toolkit for inferring adaptive lymphocyte clonality from single-cell transcriptomics lacking antigen receptor sequencing data (**Fig. 3a**). Inspired by approaches used in clinical diagnostics of B and T cell malignancies, cell2clone takes advantage of mutually exclusive expression of immunoglobulin light chain genes (*IGKC*/*IGLC*) in single B cells^29^, and T cell receptor constant chain genes (*TRBC1*/*TRBC2*) in single T cells^30,31^. Each B or T cell is classified into kappa/lambda or TRBC1/TRBC2 classes to infer clonal restriction within phenotypically defined cell populations. Non-clonal cells, such as naive B or T cells showed constant proportions of the two classes across individuals, whereas cell types containing clonal expansions exhibit skewing of the fraction to either class in individuals harbouring clonal expansions, as demonstrated using paired single-cell RNA sequencing (scRNA-seq) and antigen receptor sequencing datasets (**Extended Data Fig. 4**).

**Figure 3.**
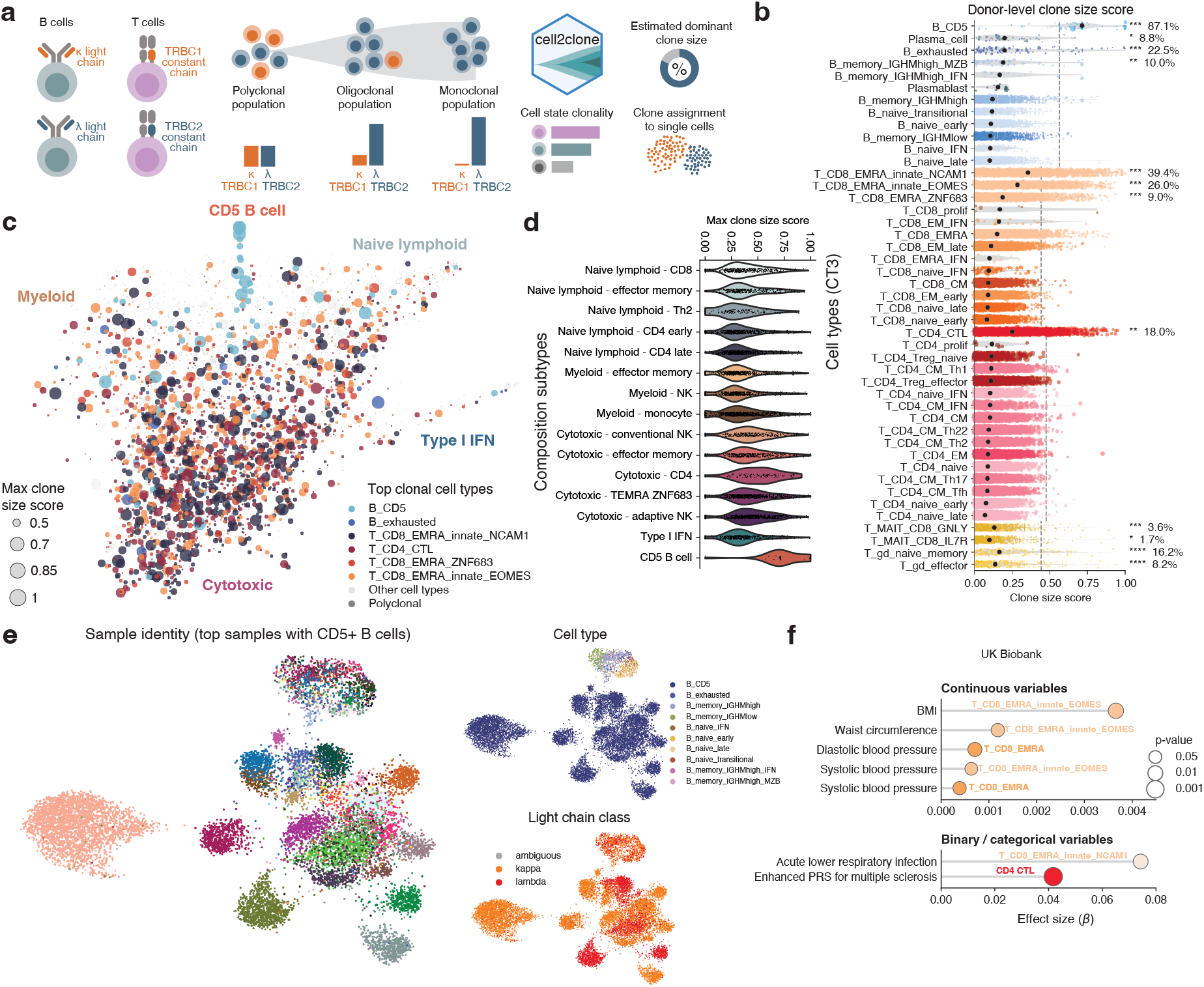
Lymphoid clonal expansions across blood archetypes and cell states. **a,** Schematic of the cell2clone framework for inferring cell state-specific clonality in adaptive lymphocytes. **b,** Inferred dominant clone size across samples in B cell, CD8+ T cell, CD4+ T cell, MAIT and gamma-delta T cell subtypes. Dots indicate individual samples, dot size corresponds to the fraction of the cell type out of the compartment in that sample, and black dots indicate median across samples. Only donors with at least 20 cells of each cell type are included in the analysis. Asterisks indicate significance based on the permutation test assessing presence of high-clonality samples in the cell type compared to non-clonal naive cells: **** FDR<0.0001, *** FDR<0.001, ** FDR<0.01, * FDR<0.05. Percentages indicate samples above the threshold of clone size score defined based on non-clonal naive cells (99.9th percentile), indicated with a dashed line. **c,** Blood composition map on a force-directed layout showing the largest clonally expanded lymphocyte state as dot colour and estimated clone size as dot size for each sample. **d,** Inferred clone size of the largest clonally expanded lymphocyte state for each sample, split by blood composition subtype. **e,** UMAPs showing all B cells from samples with the highest clone size scores and fractions of CD5+ B cells, coloured by sample, cell type and inferred light chain class. **f,** Dot plot showing the most significant features correlating with cell type-specific clonality in T cells in UKB.

Clonal expansion varied markedly across immune cell states. CD5+ B cells displayed the highest degree of clonality, consistent with MBL^18^ (**Fig. 3b**). Additionally, exhausted B cells, corresponding to the broader family of atypical or age-associated B cells^32,33^, showed extreme clonal expansions in a subset of samples, indicating that previously unappreciated clonal expansions may explain variation in the abundance of this B cell subset across individuals (**Extended Data Fig. 5g**). Similarly, IGHM^high^ memory B cells and plasma cells showed increased clonality, consistent with their antigen-experienced states. In T cells, CD8+ terminally differentiated effector memory (TEMRA) subsets, particularly innate-like CD56+ TEMRA cells, and CD4+ CTLs showed markedly increased clonality. Generally, clonality showed only modest overlap between different cell types across samples, apart from NCAM1^high^ and EOMES^high^ innate-like CD8+ TEMRA subsets, indicating likely distinct drivers of clonal expansion between different cell states and underscoring the need for cell state-aware clonality inference (**Extended Data Fig. 5a-c**).

This approach allowed us to define the landscape of adaptive lymphocyte clonal expansions across the two biobank cohorts and blood archetypes (**Fig. 3c**). In addition to the CD5+ B cell archetype, the cytotoxic archetype showed significantly increased clonality, suggesting that clonal expansion contributes to composition changes that define these archetypes (**Fig. 3d**).

To determine the phenotypic consequences of clonal expansion within blood archetypes, we examined cells from samples harbouring extreme clonal expansions at single-cell resolution, using cell2clone to assign single cells to putative clones based on transcriptomic similarity and shared antigen receptor chain class. CD5+ B cells constituted highly sample-specific clusters, while conventional B cells from different samples were mixed among clusters driven by cell state identity (**Fig. 3e, Extended Data Fig. 5d).** We observed similar relationships across other cell types, such as exhausted B cells, in high-clonality samples but not in the same cell types in low-clonality samples (**Extended Data Fig. 5e-f**), suggesting that clonal expansion can steer cells toward diversified phenotypic extremes^34^.

Across UKB metadata, clonality was associated with distinct health record variables depending on cell type (**Fig. 3f**). Clonality of innate-like CD56+ TEMRA cells correlated with lower respiratory tract infections, while clonality of EOMES^high^ innate-like CD8+ TEMRA cells correlated with higher body mass index and elevated blood pressure, consistent with the cytotoxic archetype being associated with these risk factors. CD4+ CTL clonality was associated with increased polygenic risk for multiple sclerosis, potentially linking specific clonal immune states to autoimmune predisposition.

Taken together, cell2clone revealed the landscape of cell type-specific clonal expansions across population-scale blood cohorts, identifying clonal expansions as key characteristics of the cytotoxic and CD5+ B cell archetypes and previously unappreciated clonal expansions in exhausted B cells, with potential clinical implications for recognising clonal immune dysregulation in chronic disease and interpreting blood abnormalities.

### Chromosomal alterations in CD5+ B cell archetype

Clonal expansion of blood cell states may be driven by somatic genetic alterations. We therefore asked whether somatic alterations reshape blood archetypes and contribute to progression toward extreme blood states. In particular, mosaic chromosomal alterations (mCAs) have been identified using bulk DNA sequencing across population cohorts^11,35,36^, but the cell types carrying them or their functional consequences remain poorly understood. We reasoned that highly clonal blood cell populations would provide the greatest power to detect somatic copy number alterations (CNAs) from single-cell transcriptomics and therefore focused analyses on clonally expanded B and T cell states using inferCNVpy^37^ (**Fig. 4a**).

**Figure 4.**
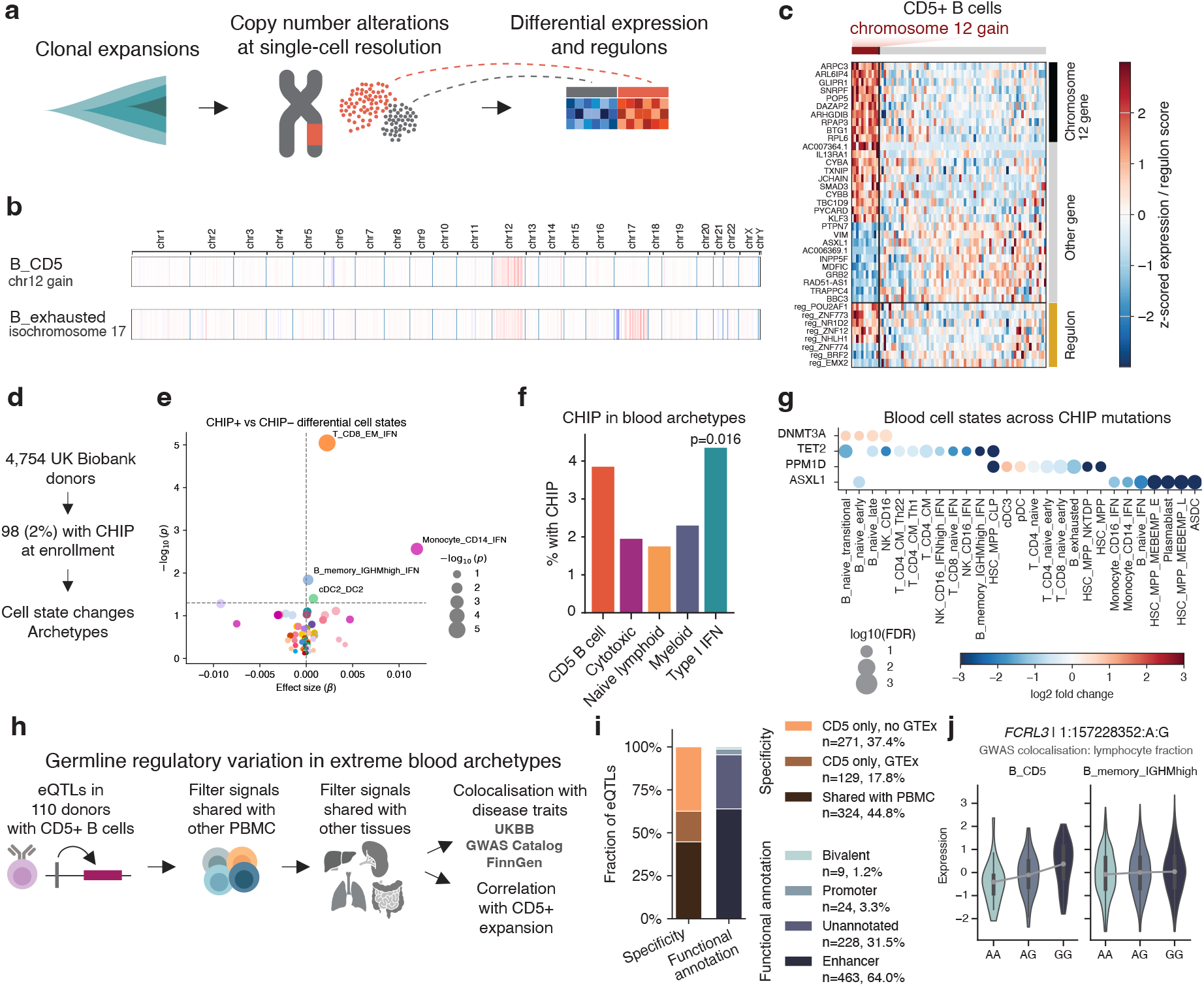
Somatic and germline genetic alterations shape cell states across blood archetypes. **a,** Workflow of identifying copy-number alterations and their consequences in clonally expanded cells. **b,** Aggregate chromosome heatmaps of CD5+ and exhausted B cells from samples carrying chromosomal alterations. **c,** Heatmap showing significantly differentially expressed genes and differentially active regulons in CD5+ B cells from donors with or without a chromosome 12 gain. **d,** Workflow of identifying associations of CHIP with cell states and archetypes. **e,** Volcano plot showing differentially enriched cell types between donors harbouring CHIP and those without, adjusted for age, sex and ancestry. **f,** Barplot showing fraction of donors with CHIP in each archetype. Significant p values from a Fisher’s exact test are shown. **g,** Dot plot showing differentially enriched cell states between each CHIP mutation and all other mutations. **ih** Workflow for identifying germline regulatory variation specific to a rare blood archetype and clonally expanded cell state (CD5+ B cells). **ji** Bar plot showing proportions of CD5+ B cell eQTLs based on cell type/tissue specificity and functional annotations. **j,** Example of dosage effect of an eQTL specific to CD5+ B cells in comparison with other B cell types, involving *FCRL3* influencing B cell receptor signalling and colocalising with lymphocyte fraction in GWAS.

In CD5+ B cells, recurrent chromosome 12 gains represented the most frequent alteration (11/78 clonally expanded samples, 14.1%), linking previously identified bulk mCAs to clonally expanded CD5+ B cells (**Fig. 4b, Extended Data Fig. 7a**). Exhausted B cells showed recurrent 17p losses coupled with 17q gains, suggesting isochromosome 17 (i(17q)) alterations (10.7% of samples with significant clonal expansions), implicating exhausted or atypical B cells in mCA-driven somatic evolution. In clonal T cell expansions, mCAs were comparatively rare, with putative gains involving chromosome 22q gains and chromosome X in EOMES^high^ innate-like CD8+ TEMRA cells NCAM1^high^ innate-like CD8+ TEMRA cells.

To define the molecular consequences of mCAs, we compared transcriptional programmes between cells carrying specific mCAs and matched cells from samples without these alterations (**Supplementary Table 9**). In CD5+ B cells harbouring chromosome 12 gains, expression changes extended beyond chromosome 12 itself, including increased expression of *KLF3* and *IL13RA1* promoting B cell development and transformation^38–40^, as well as decreased expression of *PTPN7*, a suppressor of MAPK signalling (**Fig. 4c, Extended Data Fig. 6a-b**). Activity of the B cell differentiation and malignancy-associated^41,42^ POU2AF1 (BOB-1) and EBF1 regulons were increased from the initially downregulated activity in CD5+ B cells compared to normal B cells, suggesting a partial reversion to a more conventional B cell state. In exhausted B cells, i(17q) alterations were associated with decreased *TP53* expression and increased STAT3 regulon activity, consistent with *TP53* being located in chromosome 17p and *STAT3* in chromosome 17q (**Extended Data Fig. 6c-e**). Moreover, genes such as *CCR7* were upregulated in exhausted B cells with i(17q), suggesting potential surface markers for early detection of such expansions with chromosomal alterations linked to high risk.

Single-cell resolution further revealed evidence of subclonal evolution within clonal expansions. In one sample harbouring a chromosome 12 gain in CD5+ B cells, we discovered two putative clones inferred by cell2clone, one marked by kappa light chain restriction and chromosome 12 gain and the other marked by lambda light chain restriction, potentially suggesting two independent events of clonal expansion (**Extended Data Fig. 7b**). In another sample, chromosome 12 gain occurred within a subset of expanded exhausted B cells already carrying i(17q), consistent with ongoing subclonal evolution and pinpointing potential mechanisms such as *KRAS* overexpression (**Extended Data Fig. 7c**).

In conclusion, we link specific mCAs to CD5+ and exhausted atypical B cells, elucidating mechanisms of clonal evolution with potential implications for early detection and prevention of malignant progression.

### Clonal haematopoiesis across blood archetypes

Clonal haematopoiesis of indeterminate potential (CHIP), marked by somatic mutations found in myeloid malignancies could influence mature blood cell states, blood archetypes and underlie outlier blood states. Across the CARDINAL UKB cohort, we identified 98 individuals harbouring CHIP at study enrolment, most frequently involving *DNMT3A* (n=71), *TET2* (n=17), as well as *ASXL1* and *PPM1D* (**Fig. 4d**). Multiple myeloid cell types were enriched in CHIP carriers, including S100+ CD14+ monocytes and type 2 conventional dendritic cells (cDC2), whereas lymphoid states such as naive B cells were depleted (**Extended Data Fig. 6f**). However, after adjustment for age, sex and ancestry, several type I IFN states in addition to cDC2 were enriched in CHIP carriers (**Fig. 4e**). Consistently, CHIP carriers were enriched in the type I IFN archetype (**Fig. 4f**), suggesting myeloid bias to be a more general age-related trend and a link between the IFN-responsive inflammatory blood states and CHIP.

To investigate whether specific CHIP mutations have distinct effects on blood cell states, we evaluated the cell state proportions enriched in donors carrying each of the most common mutations relative to other mutations (**Fig. 4g**). Donors with *DNMT3A* mutations had higher proportions of conventional CD16+ NK cells and naive B cell subsets. Conversely, *TET2* mutations were associated with reduction of naive B cell subsets and conventional CD16+ NK cells, as well as CD4+ central memory T cells and circulating common lymphoid progenitors. Donors carrying *PPM1D* mutations had lower CD4+ naive T cells, exhausted B cells and lymphoid progenitors. *ASXL1* mutations were associated with reduced inflammatory IFN^high^ monocyte states.

Collectively, these findings suggest a link between CHIP and the type I IFN blood archetype, potentially reflecting previously reported increased infection susceptibility^43,44^ or inflammatory priming associated with CHIP^45^.

### Regulatory variants in outlier archetypes

We reasoned that extreme blood cell states generated by somatic clonal expansion may create a regulatory context in which germline variants, including disease-associated loci, exert effects on gene expression that are not detectable in normal cell types found widely across the population. The number of individuals harbouring the CD5+ monoclonal B cell lymphocytosis cell state (n=110, at least 10 cells) representing lymphoid clonal haematopoiesis allowed us to identify expression quantitative trait loci (eQTLs) specific to this archetype driven by a premalignant blood cell state (**Fig. 4h**).

We identified 724 conditionally independent eQTL association signals involving 687 distinct eGenes in CD5+ B cells (Supplementary Table 10). Of these eQTL signals, 400 (55.25%) were not shared across the blood cell types in our data and 271 (37.4%) were not shared either with GTEx non-blood tissues, indicating that these variants are highly specific in influencing gene expression only in the context of this rare blood archetype reflecting somatic evolution (**Fig. 4i**). Several identified eGenes were involved in potentially relevant biological processes, including *FCRL3* encoding a surface receptor involved in B cell signalling^46^ and CLL prognosis^47^, *PIK3CG*^48^, *AKT2*^49^, *ZBTB17*^50,51^ linked to B cell signalling and maturation, *TCL1A* linked to clonal haematopoiesis^52^ and *LPP* linked to CLL predisposition^53^ (**Fig. 4j Extended Data Fig. 8a-c**). Additionally, we identified eQTLs impacting genes that define clinical risk in CLL, including *CXCR4*^54^, suggesting that germline genetic variation acting at the premalignant stage may impact further disease progression.

Of the eQTLs exclusively identified in CD5+ cells, 37/271 colocalised with GWAS hits for diseases and quantitative traits, indicating that lymphoid clonal haematopoiesis may enable these risk variants to influence gene expression in blood. For example, variants colocalised with lymphocyte fraction GWAS involved *FCRL3* and *ATG7*, suggesting potential roles of CD5+ B cell-exclusive variants in lymphocytosis (**Fig. 4k, Extended Data Fig. 8d**). Variants suggesting links between the CD5+ archetype and common diseases included those in *GBP3* colocalising with allergic rhinitis and *WDR34* colocalising with asthma and allergic disease (**Supplementary Table 10).**

Finally, we asked if CD5+ B cell-exclusive variants influenced genes whose expression correlated with expansion of this cell state across donors. Genes including *TGFBR3* both correlated with increased fraction of CD5+ B cells and were influenced by eQTLs (**Extended Data Fig. 8e-f**). Collectively, these results identify germline variant effects restricted to clonally expanded premalignant states and suggest that some of them may be linked to establishing the CD5+ B cell archetype and progression from lymphoid clonal haematopoiesis through clinically detectable lymphocytosis towards CLL.

### Archetypes resolve blood count abnormalities

We reasoned that the blood archetypes could facilitate diagnostics of clinical blood count abnormalities by revealing subtypes reflecting the mechanisms and cell types involved, aided by composition trajectories, clonality and GRN activity inference.

We integrated complete blood counts with the cell state fractions and archetypes and defined blood count abnormalities based on clinically used cutoffs (**Fig. 5a**). Common leukocyte abnormalities, including monocytosis, lymphocytosis and lymphopenia were enriched in distinct archetypes and composition subtypes (**Fig. 5b-c, Extended Data Fig. 9a**), similarly as a broader set of abnormalities across CBC parameters (**Extended Data Fig. 9b**). For example, both monocytosis and lymphopenia were associated with two distinct composition subtypes, type I IFN and monocyte-rich myeloid. Lymphopenia and monocytosis in the type I IFN-archetype manifested in largely non-overlapping samples, suggesting that some individuals are more prone to respond to interferon by an increase in circulating monocytes, while others respond with decreased lymphocytes (**Extended Data Fig. 9c**). Lymphocytosis could be grouped into CD5+ B cell-driven, CD8+ ZNF683^high^ TEMRA-driven, CD4 CTL-driven or adaptive NK-driven subtypes, with the CD4+ CTL-driven cases exhibiting increased clonality.

**Figure 5.**
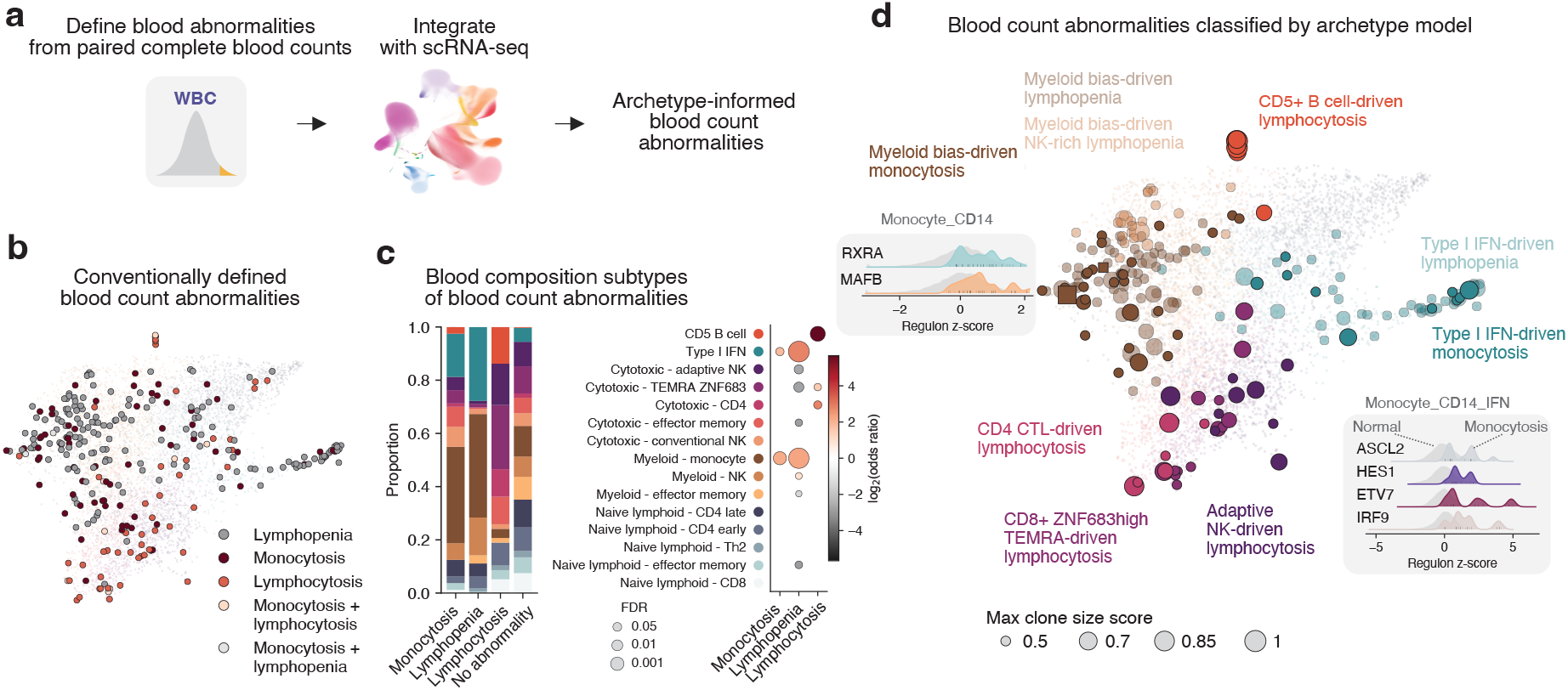
Blood archetypes resolve molecular underpinnings of clinical blood abnormalities. **a,** Workflow for integrating complete blood counts with scRNA-seq, blood composition subtypes and archetypes. **b,** Examples of clinically defined blood count abnormalities coloured on the blood composition map. **c,** Enrichment of blood composition subtypes in blood count abnormalities. Stacked bar plot shows the proportion of donors with each blood composition subtype stratified by the blood count abnormality. Dot plot shows the enrichment of the blood composition subtypes in the blood count abnormalities. **d,** Blood count abnormalities classified based on the archetype model coloured on the blood composition map. Dot size indicates inferred clone size score of the most clonal cell type in each sample. Insets show most overactive regulons in the indicated cell type between samples from the defined abnormality and control samples.

We next sought to define regulon activities underlying blood count abnormalities (**Fig. 5d**). This approach revealed two types of monocytosis, myeloid bias-driven monocytosis with heightened RXRA and MAFB regulon activity, and type I IFN-driven monocytosis with increased ASCL2, HES1, ETV7 and IRF9 activity.

We found that blood abnormalities were more common in samples with lineage sweeps than those without (**Extended Data Fig. 9d-e**). However, lineage sweeps were found also in samples with no abnormalities in CBC, indicating that the scRNA-seq-derived cell states can reveal extreme blood skewing in the absence of blood count abnormalities. Moreover, not all cases of expansive blood abnormalities such as lymphocytosis manifested as lineage sweeps in scRNA-seq, indicating utility of scRNA-seq in distinguishing phenotypically skewed expansions.

### Archetypes stratify infection and immunotherapy outcomes

Finally, to enable classification of archetypes and composition subtypes in external datasets, we developed ArcheTypist, a classification model based on the CellTypist framework^55^ (**Fig. 6a**). Following label transfer of our cell state annotations to a dataset, ArcheTypist uses the matrix of cell type fractions across samples to classify each sample into blood archetypes or more fine-grained composition subtypes, as well as to project samples to a reference embedding using nearest neighbours in the cell state fraction space. ArcheTypist achieved high performance in a held-out test set split from the CARDINAL dataset both for archetypes (F1 score 0.83-0.87) and composition subtypes (F1 score 0.57-0.85), allowing subtyping blood composition in external single-cell cohorts (**Extended Data Fig. 10**).

**Figure 6.**
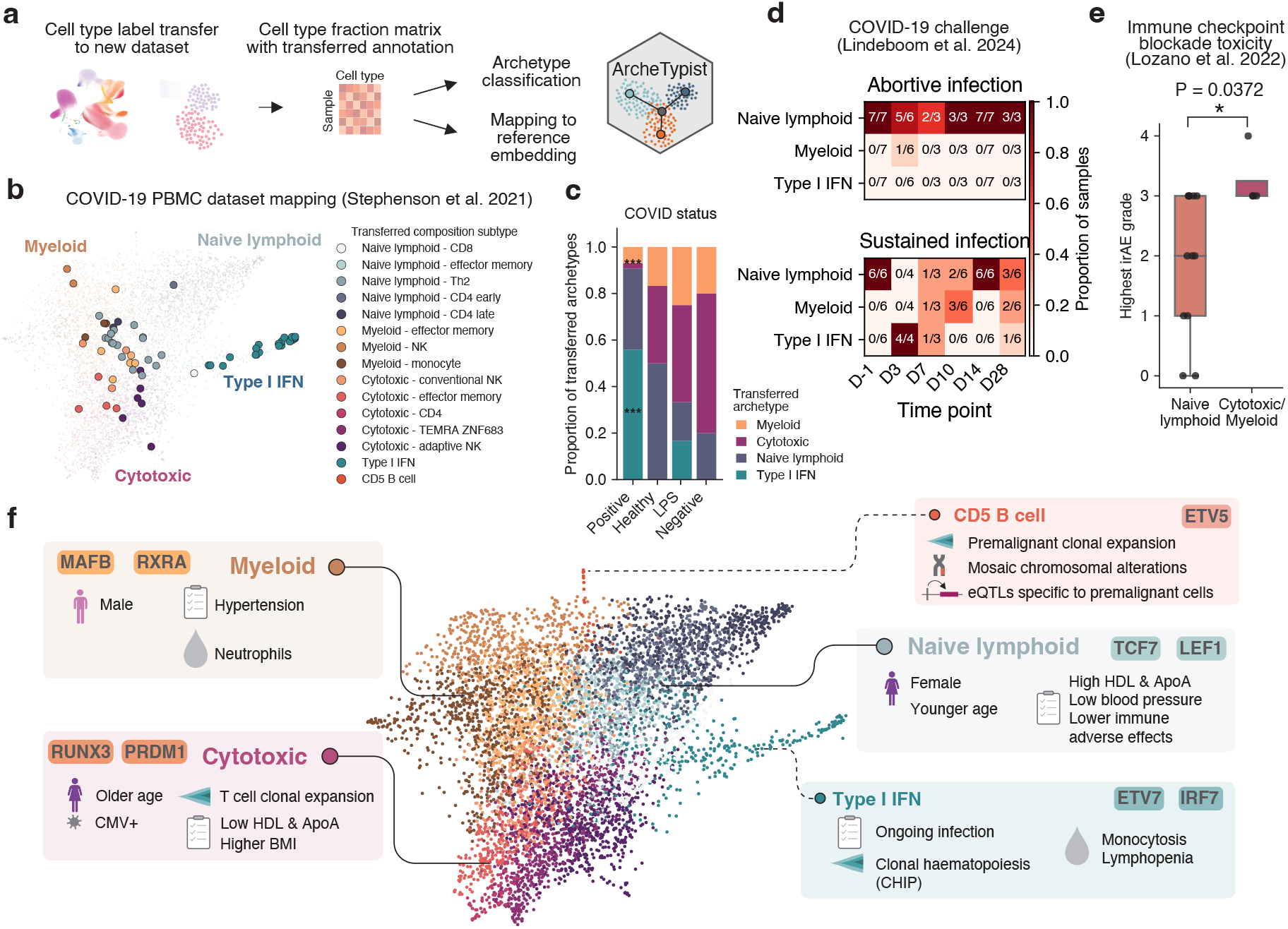
Mapping samples to blood archetypes predicts infection and immunotherapy outcomes. **a,** Workflow for classifying samples from external datasets into archetypes and blood composition subtypes and for mapping them to the reference embedding. **b,** Samples from a COVID-19 PBMC study (Stephenson et al. 2021) mapped onto the reference blood composition map with the transferred blood composition subtypes coloured. **c,** Association of archetypes with COVID-19 swab results of samples from the COVID-19 PBMC study (Stephenson et al. 2021). Stacked bar plot shows the proportion of each archetype stratified by swab result status, with significantly enriched (type I IFN) or depleted (cytotoxic) archetypes marked by asterisks. **d,** Blood archetypes from COVID-19 human challenge (Lindeboom et al. 2024) across time points stratified by type of infection (abortive/sustained). Heatmap colour and values indicate the fraction of donors in the indicated archetype out of total donors in the infection type group. **e,** Immune-related adverse events (irAE) in immune checkpoint blockade-treated melanoma patients stratified by pretreatment blood archetype. Box plot shows the highest grade of irAE for each patient shown as dots. P value is obtained using Mann-Whitney U test between naive lymphoid and other (cytotoxic/myeloid) archetypes. **f,** Summary schematic of insights into blood variation across the population revealed by the archetype model.

Demonstrating the utility of mapping external samples to our reference atlas and archetypes, blood samples from COVID-19 positive donors^4^ were enriched in the type I IFN archetype, particularly within a short duration since symptom onset, and depleted in the cytotoxic archetype (**Fig. 6b-c, Extended Data Fig. 9f-g**). In longitudinal blood samples from a COVID-19 human challenge study^5^, donors who remained stably in the naive lymphoid archetype experienced abortive or transient infections, whereas progression through the type I IFN archetype to the myeloid archetype was associated with sustained infection (**Fig. 6d, Extended Data Fig. 9h**). In pre-treatment blood samples from a cohort of anti-PD-1/anti-CTLA-4 checkpoint blockade cancer immunotherapy-treated melanoma patients^56^, the naive lymphoid archetype was associated with lower immune-related adverse events compared to the cytotoxic and myeloid archetypes (**Fig. 6e**). Taken together, these results show that blood archetypes provide a framework for translating variation in blood immune composition into clinically meaningful patient stratification across disease contexts.

## Discussion

Based on thousands of blood transcriptomic profiles at cellular resolution, we propose that blood states can be grouped into archetypes comprising individuals with similar blood composition. We use this framework to contextualise axes of variation in human blood across the population, including phenotypic extremes and clonal expansion, and ultimately to explain variation in blood counts and disease-associated perturbations (**Fig. 6f**).

The archetype model enables placing individuals along trajectories towards increasingly extreme blood compositions. Originating from a CD8+ naive lymphoid composition dominating in young individuals, the blood composition of an individual may progress towards a naive CD4+ composition more common in females, a cytotoxic archetype associated with CMV-induced clonal expansion of cytotoxic lymphocytes^8^ more common over age, or a myeloid archetype more frequent in males. In addition to demographics, blood archetypes were associated with risk factors for common diseases. The naive lymphoid archetype was linked to a more favourable cardiometabolic profile and may represent a healthier immune state. In contrast, the myeloid archetype may reflect ageing-associated myeloid bias^21,22^, potentially exacerbated by cardiovascular risk factors. By resolving coordinated variation across fine-grained cell states, blood archetypes may explain the cellular basis of earlier stratifications based on flow cytometry or bulk transcriptomics^57,58^ and of coarse metrics such as the neutrophil-to-lymphocyte ratio, which predicts risk in cardiovascular disease^59^ and cancer^60^. Progression towards archetypal extremes was accompanied by lineage sweep, a progressive reduction in blood compositional diversity driven by expansion of restricted cellular programmes, representing an important and previously underappreciated dimension of immune variation. We identify three broad routes towards such extremes. In the cytotoxic archetype, extreme states arise through adaptive clonal expansion of cytotoxic lymphocytes, consistent with chronic antigen exposure such as CMV⁸. In the CD5+ B cell archetype, they reflect premalignant lymphoid expansion accompanied by regulatory rewiring away from conventional B cell programmes, chromosomal alterations that link bulk mCA findings^11,35,36^ to a specific cell state, and germline eQTLs detectable only within this premalignant state and not in common cell types. The eQTL finding suggests that somatic evolution can unmask latent germline regulatory effects. Myeloid clonal haematopoiesis, in turn, was associated with the type I IFN archetype, which may reflect the increased infection susceptibility^43,44^ or inflammatory priming^45^ reported in CHIP carriers. Defining the regulatory programmes that sustain these outlier states may reveal ways to prevent progression towards dysfunctional ageing-associated states or malignancy.

We provide the cell2clone framework for studying clonality in emerging population-scale immune atlases^2,3,61–65^. This approach allowed us to link clonal expansion to lymphocyte subsets beyond those previously known to harbour clonal expansions. Clonal expansions in exhausted or atypical B cells raise the possibility that they may underlie monoclonal B cell lymphocytosis with a non-CLL CD5-phenotype, or represent the cell of origin of rare B cell malignancies carrying the i(17q) aberration^66,67^. We envision that cell2clone unlocks a new dimension of biology in population-scale immune atlases by enabling analyses of genetic determinants of clonal expansion across diverse cohorts and guiding research into lymphoid clonal haematopoiesis.

Clinically, the archetype framework resolves blood count abnormalities that appear indistinguishable in CBC into biologically distinct subtypes. ArcheTypist places samples from focused studies in a population-scale context, in an analogous manner to how cellular reference atlases allow placing single cells in the context of cell type and state variation. ArcheTypist linked progression through the type I IFN archetype to the myeloid archetype to sustained COVID-19 infection, and the naive lymphoid archetype to lower immune-related adverse events after checkpoint blockade in melanoma patients. These results point towards a next generation blood diagnostic measurement, CBC 2.0, integrating blood archetypes, cell states, clonality, regulatory programmes and somatic alterations.

Our study has limitations. The cross-sectional nature of the data prevents assessing whether a person’s archetype is stable over time, warranting future longitudinal sampling. Moreover, the direction of causality of archetype associations with health records remains undefined. In addition, clonality and copy-number alterations inference from transcriptomes captures mainly large clones, potentially underestimating their frequencies.

Population-scale single-cell atlases make it possible to define the normal range of tissue composition, the recurrent subtypes within it, and the outliers at its edges. The archetype framework offers a general model for this task. Beyond blood, applying the same principles to emerging population-scale datasets of solid tissues can enable identification of tissue archetypes, trajectories, and pathological extremes across diverse human organs.

## Methods

### The CARDINAL dataset and cell annotations

The CARDINAL scRNA-seq dataset generation, including sample collection, preprocessing and cell annotation strategy including marker genes are described in Giacopuzzi et al.^17^ Complete blood counts from the same samples were obtained using a Sysmex XN-1000 haematology analyser as described in Giacopuzzi et al.^17^

### Defining blood composition subtypes and archetypes

To assign samples into subgroups based on the composition of co-varying cell types, a sample-by-cell-type matrix with each feature representing the proportion of a given cell type (CT3 level) within the cells in the sample was generated. For easy exploration and analysis, the cell composition atlas was loaded into an AnnData^68^ object, with cell types as “var_names”, samples as “obs_names” and cell type fractions as the data matrix, enabling analysis using Scanpy^69^ in a similar manner as a conventional single-cell object. Convenience functions to facilitate object building are available as part of our companion package ArcheTypist. Principal component analysis (PCA) was performed and neighbours and graph-based Leiden clusters of samples reflecting similar cell type compositions (*blood composition subtypes*) were defined using Scanpy with default parameters. To identify characteristic cell types for the archetypes, differences between cell type composition in each blood composition subtype compared to all others were tested using Mann-Whitney U-test on CT3 fractions out of total cells per donor, followed by Benjamini-Hochberg adjustment for multiple testing. Cell composition clusters enriched in similar cell types were merged to obtain blood subtypes termed *archetypes*. The archetypes and blood composition subtypes were visualised on a force-directed atlas embedding computed using the neighbours in the cell composition space using “sc.tl.draw_graph” in Scanpy.

### Associations with biobank metadata

Associations between metrics derived from scRNA-seq, including archetypes, blood composition subtypes, cell state entropy and clonality, and UK Biobank clinical phenotypes were evaluated using sample-level measures treated as binary variables (archetype and blood composition subtype membership) or continuous variables (cell state entropy and clonality). Diagnosis variables defined by recorded dates (for example, first reported disease events) were converted into binary “ever” indicators, with non-missing dates coded as cases. Continuous phenotypes were analysed directly after numeric coercion. Variables with insufficient case counts or limited non-missing observations were excluded to ensure model stability.

For each immune metric–phenotype pair, we fitted linear regression models with the immune metric as the dependent variable and the phenotype of interest as the independent variable. Models were adjusted for age, sex, and ethnic background where indicated. Categorical covariates were encoded using reference-level dummy variables. Observations with missing values in the outcome, predictor, or covariates were excluded on a per-model basis.

Associations between archetypes or blood composition subtypes and disease in G&H were tested using generalised additive models. Separate binomial models were fitted for each archetype or composition subtype, with membership in the selected subtype or archetype coded as the binary outcome and all remaining groups as the reference. Disease status was included as the predictor of interest, with adjustment for sex, ancestry, and age. Age was modelled as a smooth term using cubic regression splines, with sex-specific smooths where specified. Models were fitted using restricted maximum likelihood. Multiple-testing correction was performed using the Benjamini–Hochberg procedure.

### Gene regulatory network inference

For GRN inference using SCENIC^25^, the UKB cohort was first subsampled to ∼280,000 cells, with including 20,000 cells from medium-abundance cell types (at least 1,000 cells in total) and preserving 5% of cells from abundant cell types (at least 20,000 cells in total) and 50% of cells from sparse cell types (less than 20,000 cells in total) at the CT3 level. Cell-type-specific regulons were inferred using pySCENIC^70^, which infers TF–target gene relationships based on gene co-expression and enrichment of TF-binding motifs near target gene promoters. To reduce the impact of sparsity in single-cell expression data and improve computational tractability, cells were further aggregated into mini-metacells comprising 10–15 cells. After obtaining the regulons from SCENIC, the regulons were scored using “[sc.tl](http://scanpy.tl/).score_genes” in Scanpy and an object of patient-specific pseudobulks at the CT3 level comprising both the UKB and G&H cohorts was obtained. Regulons differentially active between cell types were identified using a Mann-Whitney U test between pseudobulks of one cell type and comparator cell types, requiring at least 20 cells for a pseudobulk to be included in the analysis. Regulons were correlated with cell type fraction out of all PBMCs using Spearman correlation, requiring at least 20 cells for a pseudobulk to be included in the analysis.

### Cell composition trajectory inference

Cell composition trajectories were inferred using scFates. The cell type fraction-based PCA and neighbours was used as basis and the tree computed using function scf.tl.tree in scFates with the following parameters: method=“ppt”, Nodes=125, ppt_lambda=100, ppt_sigma=0.0125, ppt_nsteps=100. Results were projected on the force-directed atlas embedding. A root was selected based on the vicinity of a trajectory branch terminal node to the CD8+ naive lymphoid composition subtype, identified as the composition subtype highly enriched in young individuals. Pseudotime was computed using [scf.tl](http://scfates.tl/).pseudotime in scFates. Milestones were annotated based on their overlap with previously defined cell composition subtypes. Cell type fractions were correlated with pseudotime along a trajectory between a starting point, such as the naive lymphoid CD8+ milestone, and other terminal milestones using Spearman correlation.

### Cell composition entropy

Shannon entropy was used to quantify the diversity of cell state composition within each donor. Cell state fractions were first averaged across all samples from the same donor and normalised so that fractions summed to one. Shannon entropy was then calculated using “scipy.stats.entropy” in SciPy^71^ on the resulting relative cell state distribution using a base-2 logarithm. To avoid undefined values arising from zero fractions, a small pseudocount (10⁻¹²) was applied before computation. Higher entropy values indicate a more even distribution of cell states, whereas lower values indicate dominance by a smaller number of cell states. Cell state entropy was calculated after subsampling each donor to 250 cells. Donors with < 250 total cells were excluded from the analysis, as entropy was found markedly reduced in these donors likely due to insufficient sampling. Lineage sweep was defined as cell state entropy < 4.5.

### Adaptive lymphocyte clonality inference using cell2clone

We built the cell2clone workflow for identifying clonally expanded adaptive lymphocyte populations in scRNA-seq data without VDJ sequencing information. For each cell, expression of constant region genes was quantified as normalised expression values. For T cells, β-chain constant region usage was assessed using TRBC1 and TRBC2. For B cells, light chain usage was assessed using the κ-chain gene (IGKC) and λ-chain genes (IGLC), in which case normalised expression values were summed across all λ-chain genes. A cell was classified as κ (or TRBC1) if transcripts from the corresponding gene or gene set were higher than those of λ (or TRBC2) and vice versa. Cells in which neither gene set was detected were classified as ambiguous.

Clonality was quantified at the sample-by-cell-type level. We ran cell2clone across all B and T cell subtypes at the CT3 cell type level for all samples and ranked cell types by their median dominant clone size. For each sample and each annotated cell type, the number of cells classified into each state was counted after excluding ambiguous cells. Sample–cell type combinations with fewer than 20 classified cells were excluded from downstream analysis to reduce instability due to low counts. Clonality was summarised using several complementary measures, including the fraction of cells assigned to the first state and Shannon entropy as a measure of diversity within the two-state system. In addition, a median-centered directional log2 ratio of the counts of the two states was calculated to capture both magnitude and direction of imbalance, as well as absolute value of this as a directionless clonality metric. A clone size score was calculated to quantify the deviation of the largest clone fraction of each cell type from the expected value in a fully polyclonal cell population. The fraction assigned to the dominant clone was first constrained to lie within a small interval away from 0 and 1 to improve numerical stability. The expected clone fraction was then estimated either separately within each cell type (default) as the median largest clone fraction of that cell type, or globally across all cells as the overall median. For each cell type, the absolute deviation from the expected value was computed and normalised by the maximum possible deviation given the expected fraction, yielding a score bounded between 0 and 1. A score of 0 indicates that the observed clone fraction equals the expected value (i.e. fully polyclonal population), whereas a score of 1 corresponds to the largest possible deviation (i.e. fully monoclonal population).

To contextualise clonality within overall cellular composition, the proportion of the total sequenced cells in each sample represented by a given cell type was calculated using all cells from that sample, including ambiguous and unclassified cells. This separation ensured that clonality metrics reflected confidently classified cells, while abundance metrics reflected overall representation within the sampled cellular compartment.

To analyse whether clonal expansions in different cell types co-occur in the same individuals, graph-based clusters were derived based on the cell type-specific clone size score metric using Leiden clustering and the variation was visualised using a UMAP (Uniform Manifold Approximation and Projection) embedding.

### Permutation test for cell types harbouring clonal expansions

To identify cell types enriched for samples with unusually large clone size scores, a permutation-based enrichment test was performed. A reference cell type, such as naive B or T cells, was used to define a clone size score threshold corresponding to a specified percentile of its score distribution (99th percentile unless otherwise stated). For each test cell type, the observed proportion of samples with clone size scores exceeding this threshold was calculated. Statistical significance was assessed using a permutation test in which cell type labels were randomly permuted while clone size scores were held fixed, preserving the overall score distribution. For each permutation, the proportion of samples assigned to the test cell type that exceeded the reference threshold was recalculated, generating a null distribution of expected proportions under random assignment. One-sided empirical p values were computed as the proportion of permutations producing an equal or greater fraction of high-scoring cells than observed. Cell types represented in fewer than 10 samples were excluded from analysis. P values were adjusted for multiple testing across cell types using the Benjamini–Hochberg false discovery rate procedure.

### Assignment of single cells to putative clones

Clonal assignment was performed using a sample-aware clustering approach combined with receptor chain dominance criteria. Briefly, using a set of selected samples and cells restricted to an annotation hierarchy level (e.g. CT1=”B” for B cells), PCA dimensionality reduction and k-neighbour graph construction were carried out using Scanpy, with T cell receptor and immunoglobulin genes excluded from highly variable gene selection to prevent clustering from being driven by antigen receptor expression. To avoid cross-sample aggregation, Leiden clustering was performed separately within each sample.

Cells were classified according to mutually exclusive receptor chain states (TRBC1 versus TRBC2 for T cells; κ versus λ for B cells) as described above. Within each sample-specific Leiden cluster, the proportion of classified cells belonging to each chain state was calculated. Clusters exceeding a predefined minimum size and exhibiting dominance of a single chain state above a selected threshold (≥80% as default) were designated as putative clones; all other clusters were labeled polyclonal. Cells without confident chain assignment were excluded from purity calculations but retained in final labeling. This strategy identifies transcriptionally coherent, chain-dominant populations consistent with clonal expansion.

### Sample-specific transcriptional segregation analysis

To assess whether clonal expansion is associated with unique cell states restricted to samples harbouring them, sample-specific segregation of transcriptional states was quantified within each cell type using silhouette analysis in the principal component (PC) space. Analyses were restricted to samples classified as having high clonality (“top”) or low clonality (“bottom”), and to the relevant annotation hierarchy level (e.g. CT1=”B” for B cells). For each cell type and clonality group separately, cells were represented using the first 30 PCs from the gene expression PCA embedding. Samples represented by fewer than 10 cells within a given cell type were excluded. To reduce the influence of unequal cell recovery between samples, a maximum of 300 cells was randomly selected per sample.

Sample identity was used as the cluster label, and a silhouette coefficient was calculated for each cell using Euclidean distances in PC space. For a given cell (i), the silhouette coefficient was defined as (s(i)=[b(i)-a(i)]/\max[a(i),b(i)]), where (a(i)) is the mean distance between the cell and other cells from the same sample and (b(i)) is the minimum mean distance between the cell and cells from any other sample. Positive values therefore indicate preferential proximity to cells from the same sample, values near zero indicate inter-sample mixing, and negative values indicate greater similarity to cells from another sample. Cell-level silhouette coefficients were averaged within each sample to obtain one sample-level segregation score for each cell type.

For each cell type, sample-level silhouette scores were compared between the high- and low-clonality groups using a two-sided Mann–Whitney U test. Multiple testing across cell types was controlled using the Benjamini–Hochberg procedure. Higher silhouette scores in the high-clonality group were interpreted as increased sample-specific segregation of transcriptional states, whereas scores near zero indicated greater mixing of cells between samples.

### Benchmarking and validation

To benchmark cell2clone, we used a collection of six COVID-19 PBMC datasets^4,5,72–75^ with joint scRNA-seq and VDJ-sequencing data and harmonised T cell annotations compiled as part of a larger human T cell infection atlas (Dratva et al. unpublished; tcellatlas.org). Cell2clone was first run to obtain clone size scores for each cell type in each sample. Cell types containing significantly expanded clones were defined using naive CD4+ T cells as the reference with 97.5th percentile of the clone size score as threshold. Clone size scores in these cell types were then correlated with the VDJ-seq-derived fraction of cells comprising the largest clone in the matching cell type across samples using Pearson correlation. Moreover, the significance of largest clone fraction difference within these cell types between clonal and non-clonal samples as inferred by cell2clone, dichotomised for each cell type using a threshold of clone size scores based on a threshold defined by non-clonal cell types, was determined using Mann-Whitney U test.

To validate assignment of single cells to clones using cell2clone in B cells, a COVID-19 PBMC dataset^4^ containing B cells from patients with moderate severity infection with paired scRNA-seq and VDJ-seq was used. Cell2clone was run on the B cells using functions “c2c.classify_lightchain_or_trbc” and “c2c.compute_sample_celltype_clonality” with min_cells = 40 to obtain clone size score for each cell type in each sample, and samples and cell types containing significantly expanded clones were defined using naive B cells as the reference using the function “c2c.permutation_test” with 0.975 as threshold. The sample with largest clonal expansion, occurring in exhausted B cells, was examined at single-cell resolution and cells assigned to clones using the “c2c.leiden_chain_clones_sampleaware“ function. Overlap of VDJ-seq-derived and cell2clone-derived clone assignment was compared to demonstrate cell2clone performance at single-cell resolution.

### Analysis of chromosomal alterations

We used inferCNVpy to identify CNAs. To identify samples harbouring mCAs in clonally expanded cells, we focused the analysis on cell types harbouring clonal expansions based on cell2clone inference: B_CD5, B_exhausted, T_CD4_CTL, T_CD8_TEMRA_innate_NCAM1, T_CD8_TEMRA_innate_EOMES, T_CD8_TEMRA_ZNF683 and T_CD8_TEMRA. For B_CD5, we performed the analysis using all samples with at least 10 B_CD5 detected due to the uniform clonality of this cell type across samples. For B_exhausted, we focused on samples inferred by cell2clone to be more clonal than naive B cells (B_naive_early), for T_CD4_CTL samples more clonal than naive CD4+ T cells (T_CD4_naive_early) and for T_CD8_TEMRA_innate_NCAM1, T_CD8_TEMRA_innate_EOMES and T_CD8_TEMRA_ZNF683 and T_CD8_TEMRA samples more clonal than naive CD8+ T cells (T_CD8_naive_early), all at a threshold of 0.999 defined using the naive reference population.

InferCNVpy was run across all selected samples using an object containing only the cell type of interest, using average of all cells as reference for CNA inference. Samples harbouring copy number alterations with high confidence were identified by first removing samples with highly noisy CNA calls and then calling CNAs in samples using chromosome-specific thresholds for inferred log2 copy number ratio and for the fraction of cells harbouring a CNA with the selected threshold.

For samples with detected CNAs, a sample-wise InferCNVpy analysis using all B and plasma cells from each sample was performed, with B_naive_early, B_naive_late, B_memory_IGHMhigh, B_memory_IGHMlow as reference cell types. A principal component analysis was performed, neighbours were defined and Leiden clustering used in the copy number space to identify putative subclones. Subclones were annotated by examining chromosome-wise copy numbers split by copy number-based clusters.

Differentially expressed genes between samples with and without a specific CNA were defined using a Mann-Whitney U test between sample-level pseudobulks of a given cell type, followed by Benjamini-Hochberg adjustment for multiple testing. Differentially expressed genes were stratified based on the chromosome they are located in to identify CNA-associated transcriptomic alterations beyond the dosage effect driven by the CNA. Differentially active regulons were identified by first scoring regulons previously defined using SCENIC on single cells of the object containing only a single cell type from samples with clonal expansions, and then using a Mann-Whitney U test between sample-level pseudobulks of a given cell type, followed by Benjamini-Hochberg adjustment for multiple testing.

### Analysis of cell states associated with clonal haematopoiesis of indeterminate potential

Samples with clonal haematopoiesis of indeterminate potential (CHIP) were defined using the UKB data field 30105 “Clonal haematopoiesis of indeterminate potential (CHIP) number of variants” as having at least 1 CHIP mutation.

Associations between CHIP status and individual cell states were assessed using both univariable and multivariable analyses. Initially, each cell state was compared between CHIP-positive and CHIP-negative individuals. Group means were calculated and effect sizes were summarised as log2 fold changes, calculated from the ratio of group means after the addition of a small constant (10⁻⁹) to avoid division by zero. Statistical significance was evaluated using two-sided Welch’s t-tests. P values were corrected for multiple testing across all cell states using the Benjamini–Hochberg false discovery rate (FDR) procedure.

To adjust for demographic factors, a separate ordinary least squares linear regression model was fitted for each cell state. Cell state abundance was modelled as the dependent variable, with CHIP status included as the primary predictor and age, sex, and ethnic background included as covariates. For each model, the regression coefficient (β), standard error, t statistic, 95% confidence interval, and P value corresponding to CHIP status were extracted. Standardised regression coefficients were calculated by scaling β according to the standard deviations of the outcome and CHIP status. Effect sizes were additionally expressed as the percentage change relative to the overall mean abundance of each cell state. P values from all regression models were adjusted for multiple testing using the Benjamini–Hochberg FDR procedure.

Gene-specific analyses were performed in an analogous manner. For each recurrently mutated CHIP gene, individuals carrying a mutation in the gene of interest were compared with other mutation carriers. For each cell state, group means, medians, and log2 fold changes were calculated, and statistical significance was assessed using two-sided Welch’s t-tests. P values were adjusted separately for each gene across all tested cell states using the Benjamini–Hochberg FDR procedure.

### Identification of CD5+ B cell-specific cis-eQTLs in UKB

We used CARDINAL *cis*-eQTL summary statistics from 4,452 UK Biobank participants of European ancestry across 23 immune cell types (Giacopuzzi et al.^17^, together with fine-mapping and colocalisation results from Pirastu et al. ^76^ Briefly, *Cis*-eQTLs were mapped using TensorQTL^77^ within ±1 Mb of each gene’s transcription start site (gene-level q ≤ 0.05). Within the FLANDERS framework, associated loci were fine-mapped using SuSiE^78^ with in-sample LD, and signals were tested for colocalisation. Full details of eQTL mapping are provided in Giacopuzzi et al.^17^ and of FLANDERS, fine-mapping and colocalisation in Pirastu et al.^76^

To identify cis-eQTL signals specific to CD5+ B cells in UKB individuals, we systematically removed signals shared with other cellular or tissue contexts using colocalisation analyses. CD5+ B cell cis-eQTL signals were first tested for colocalisation with eQTL signals from all other UKB cell types at CT2 resolution. We additionally assessed colocalisation with non-blood bulk-tissue GTEx eQTL credible sets obtained from the Open Targets Platform (v25.09)^79^. Across these analyses, signals with a posterior probability of colocalisation (PP.H4) > 0.5 in any non-target cell type or non-blood tissue were classified as shared and excluded. Signals without evidence of colocalisation in any of these contexts were retained as CD5+ B cell-specific cis-eQTL signals.

We next assessed whether these CD5+ B cell-specific cis-eQTL signals colocalised with genetic associations for a broad range of diseases and quantitative traits. Fine-mapped GWAS summary statistics were obtained from the Open Targets Platform (v25.09) for 17,641 GWAS Catalog studies and 1,248 FinnGen R12 traits. These were complemented by 46 manually curated diseases (Supplementary Table 30 in Giacopuzzi et al.^17^) and 29 haematological traits measured across the full UKB cohort. GWAS for the haematological traits were performed using REGENIE v4.1^80^ and TOPMed-imputed genotypes following a previously described protocol^81^. Colocalisation between CD5+ B cell-specific cis-eQTLs and disease or trait loci was considered supported when PP.H4 ≥ 0.75.

To determine whether these disease and trait associations were specific to the CD5+ B cell context, each associated locus was subsequently tested for colocalisation with eQTL signals from all other UKB CARDINAL cell types at CT2 resolution and with GTEx bulk-tissue eQTLs. Associations showing evidence of colocalisation (PP.H4 ≥ 0.75) in any of these non-target cellular or tissue contexts were classified as non-specific and excluded. The remaining associations therefore represented disease or trait loci colocalising specifically with CD5+ B cell cis-eQTL signals. Finally, Mendelian randomisation was performed using the Wald ratio to estimate the effect of genetically predicted CD5+ B cell gene expression on the corresponding disease and trait associations.

### Defining clinical blood count abnormalities

#### Classification of CBC abnormalities

Each CBC measurement was classified as low, within range or high using predefined reference intervals. The intervals used were 4–10 × 10^9/L for white blood cells (WBC), 4–5.4 × 10^12/L for red blood cells (RBC), 115–155 g/L for haemoglobin, 36–48% for haematocrit, 80–100 fL for mean corpuscular volume (MCV), 150–400 × 10^9/L for platelets, 2.5–7 × 10^9/L for neutrophils, 1–4 × 10^9/L for lymphocytes, 0.2–1 × 10^9/L for monocytes, 0–0.5 × 10^9/L for eosinophils, 0–0.3 × 10^9/L for basophils and 0–0.1 × 10^9/L for immature granulocytes. Red-cell distribution width (RDW) reference intervals were 39–46 fL for RDW-SD and 11.5–14.5% for RDW-CV. These classifications were additionally mapped to conventional haematological terms, including anaemia, leukopenia, leukocytosis, neutropenia, neutrophilia, lymphopenia, lymphocytosis, monocytosis, eosinophilia, basophilia, thrombocytopenia, thrombocytosis, microcytosis, macrocytosis and increased RDW.

To determine whether blood count abnormalities occupied specific regions of the blood composition landscape, donors with abnormalities were stratified by composition subtype and blood archetype. Enrichment of samples with blood abnormalities (all high and low CBC classifications) compared to samples without these abnormalities within composition subtypes or archetypes was assessed using two-sided Fisher’s exact tests followed by Benjamini–Hochberg adjustment.

To assess whether blood count abnormalities differed in their position along the blood composition trajectories or in cell state entropy, samples were grouped as having monocytosis, lymphopenia, lymphocytosis, multiple selected abnormalities or no selected abnormality and differences in blood-composition pseudotime and Shannon entropy were evaluated using two-sided Mann–Whitney U tests. Analyses were also stratified by blood archetype, comparing each abnormality with samples without the selected abnormalities within the same archetype using two-sided asymptotic Mann–Whitney U tests.

#### Clonal expansion in composition-defined blood abnormalities

To test whether composition-defined abnormalities were associated with expanded lymphocyte clones, donor-level cell-type-specific clone-size scores were extracted from the clonality matrix. For each composition-defined abnormality and each available cell type, clone-size scores were compared with those of donors with no selected abnormality using a one-sided Mann–Whitney U test, testing for increased clonality in the abnormality group. Comparisons were performed only when at least three samples were available in each group. Benjamini–Hochberg correction was applied across comparisons. For visualisation of the blood composition manifold, the maximum clone-size score across cell types was additionally calculated for each sample.

#### Regulon analysis of composition-defined blood abnormalities

To identify transcriptional programmes associated with composition-defined abnormalities, the relevant cell-type-specific regulon scores for each abnormality were compared between samples carrying the composition-defined abnormality and samples with no selected abnormality. Samples were retained only when at least 20 cells of the relevant cell type were available and at least three eligible samples in both the abnormality and reference groups were required. For each regulon, regulon overactivity was tested using a Mann–Whitney U test, applied to phenotype-matched cell populations, including CD5 B cells in CD5+ B-cell-associated lymphocytosis, CD14+ monocytes in myeloid-biased monocytosis, IFN-responsive CD14+ monocytes in type I IFN-associated monocytosis, adaptive CD16 NK cells in adaptive NK-associated lymphocytosis, ZNF683-high CD8+ TEMRA cells in ZNF683-high TEMRA-associated lymphocytosis and CD4+ CTLs in CD4+ CTL-associated lymphocytosis. For visualisation, regulon activity distributions were represented as smoothed density ridges, with the distribution across samples shown together with the corresponding distribution and individual values for donors carrying the abnormality.

### Archetype classification and sample mapping using ArcheTypist

#### Archetype classification

To enable classification of samples into the archetypes and cell composition subtypes defined here in external datasets, we developed the ArcheTypist framework. The prediction consists of two stages. First, a conventional CellTypist^55^ prediction is run on a dataset to transfer the cell state labels defined in our study. Second, an AnnData object is derived based on cell state fractions across samples. This object is then used to classify samples into archetypes and composition subtypes with two available methods, either using another prediction round based on CellTypist logistic regression using an ArcheTypist model where features are cell states instead of genes, or a nearest-neighbour method described below. In the CellTypist-based logistic regression method, the predicted labels at single-sample resolution are used as the predictions.

To assess the reproducibility of archetypes and blood composition subtypes, samples from the CARDINAL dataset with existing archetype and composition subtype annotations were randomly divided into training (80%) and test (20%) sets. ArcheTypist was trained on the training set using archetype assignments as class labels and subsequently applied to the held-out test set. Classification performance was evaluated using precision, recall, and F1 score for each archetype, together with macro-averaged and weighted-averaged summary statistics. Performance metrics were calculated using the classification_report function from the Python package scikit-learn.

#### Mapping query to reference

Query samples were mapped onto a reference force-directed FA2 graph using similarity in the cell type fraction space. Reference and query AnnData objects were first restricted to their shared cell type features. Cell type fraction matrices were extracted from both objects and converted to dense arrays where required. For each query sample, the five nearest reference samples were identified using cosine distance on the shared cell type fraction profiles. Neighbour weights were calculated as the inverse cosine distance, with a small numerical offset to avoid division by zero, and normalised to sum to one for each query sample. Query FA2 coordinates were then inferred as the weighted average of the FA2 coordinates of the corresponding nearest reference neighbours.

Mapping diagnostics were stored for each query sample, including the closest reference sample, the nearest-neighbour cosine distance, and mapping similarity, defined as one minus the cosine distance. Reference annotations for archetype and composition subtype were transferred to query samples using a weighted nearest-neighbour vote. For each annotation, neighbour weights were summed by label, and the label with the highest total weight was assigned to the query sample. The maximum summed weight was stored as the transfer confidence.

## Supplementary Tables

**Supplementary Table 1.** Differentially enriched cell types between archetypes.

**Supplementary Table 2.** Differentially enriched cell types between blood composition subtypes.

**Supplementary Table 3.** Associations of archetypes with clinical variables in the G&H cohort.

**Supplementary Table 4.** Associations of blood composition subtypes with clinical variables in the G&H cohort.

**Supplementary Table 5.** Regulon targets for each transcription factor based on gene regulatory network inference.

**Supplementary Table 6.** Differential regulon activity of each CT3 cell type compared to all other cell types.

**Supplementary Table 7.** Differential regulon activity of each CT3 cell type compared to closely related cell types.

**Supplementary Table 8.** Correlation of regulon activity with cell type fractions across samples.

**Supplementary Table 9.** Differentially expressed genes with mosaic chromosomal alterations.

**Supplementary Table 10.** eQTLs in CD5+ B cells.

## Acknowledgements

This research was funded in part by the Wellcome Trust [grant number 220540/Z/20/A], the Chan Zuckerberg Foundation [grant number 2020-224316] and by the British Heart Foundation (BHF) Cambridge Centre for Research Excellence RE/18/1/34212. Fondazione Human Technopole is supported by public funds awarded by the Italian Ministry of Economy and Finance, the Ministry of University and Research, and the Ministry of Health.

This work is supported by the project IMPACT (G.A.N.) 101201772 Funded by the European Union. Views and opinions expressed are however those of the author(s) only and do not necessarily reflect those of the European Union or grant authority (ERCEA). Neither the European Union nor the granting authority can be held responsible for them.

This research has been conducted using the UK Biobank Resource under application number [69938].

We thank Wellcome Sanger Institute scientific operations team members; Stefanie Lensing, Scott Thurston, Lesley Shirley, Sameena Iqbal, Katy Taylor, Cordelia Langford and Ian Johnston for automation, sample management and sequencing operations support. Wellcome Sanger Institute Human Genetics Department team members; Sally Bygraves, Nicola Corton, Kalia Dede, Guillaume Noel and Hannes Postingl.

Genes & Health is/has recently been core-funded by Wellcome (WT102627, WT210561), the Medical Research Council (UK) (M009017, MR/X009777/1, MR/X009920/1), Higher Education Funding Council for England Catalyst, Barts Charity (845/1796), Health Data Research UK (for London substantive site), and research delivery support from the NHS National Institute for Health Research Clinical Research Network (North Thames). We acknowledge the support of the National Institute for Health and Care Research Barts Biomedical Research Centre (NIHR203330); a delivery partnership of Barts Health NHS Trust, Queen Mary University of London, St George’s University Hospitals NHS Foundation Trust and St George’s University of London. Genes & Health is/has recently been funded by Alnylam Pharmaceuticals, Genomics PLC; and a Life Sciences Industry Consortium of AstraZeneca PLC, Bristol-Myers Squibb Company, GlaxoSmithKline Research and Development Limited, Maze Therapeutics Inc, Merck Sharp & Dohme LLC, Novo Nordisk A/S, Pfizer Inc, Takeda Development Centre Americas Inc. We thank Social Action for Health, Centre of The Cell, members of our Community Advisory Group, and staff who have recruited and collected data from volunteers.

We thank: Barts Health NHS Trust, NHS Clinical Commissioning Groups (City and Hackney, Waltham Forest, Tower Hamlets, Newham, Redbridge, Havering, Barking and Dagenham), East London NHS Foundation Trust, Bradford Teaching Hospitals NHS Foundation Trust, Public Health England, Discovery Data Service/Endeavour Health Charitable Trust (especially David Stables), Voror Health Technologies Ltd, NHS England - for GDPR-compliant data sharing backed by individual written informed consent.

Most of all we thank all of the volunteers participating in UK Biobank and Genes & Health.

## Author contributions

O.D. conceptualised the study and designed and performed analyses. S.L. contributed to study design and performed analyses. J.P.P. performed gene regulatory network inference. A.L. performed eQTL analyses. D.Z. developed biobank metadata analysis approaches. L.Dratva and L.K. contributed to clonality analysis benchmarking. K.P. contributed to computational tool development. C.X. and A.-M.C. contributed to cell type annotation. I.S. contributed gene regulatory network analysis design.

G.I., D.v.S., E.C., F. Torabi, M.H., N. Subasinghe, L.D. and S.W. generated the data.

M.O., A.R., D.B., E.G., R.E., and V.I. performed data processing.

E.G., S.W., M.O., A.C., and A.R. carried out data quality control and benchmarking.

J.P.E., S.L., A.R., O.D. and F.C. conducted data integration, cell annotation and refinement.

M.O., S.S., A.R., A.S., B.A., E.H., M.J.B. and N.P. performed eQTL analyses. O.D., G.C., J.P.E., L.A., E.R., B.Costa, M.T., P.B., D.V., B.C., F.P.C., O.S., C.A.G. and N.Soranzo analysed cellular and gene programs.

C.C., M.I.C., I.G., C.D.C., M.C., A.G., V.S., B.S. assisted with immunological interpretation.

V.S. and G.V. assisted with haematological interpretation.

J.W., R.T. and S.F. provided research resources.

V.I., F.J.T., O.S., D.A.v.H., N.Soranzo and S.A.T. supervised the study.

O.D., S.L., J.P.P., A.L., I.S., M.T., C.D.C., F.C., D.A.v.H., N.Soranzo and S.A.T. wrote the manuscript.

All authors reviewed and approved the final manuscript.

## Consortia

### Genes & Health Research Team

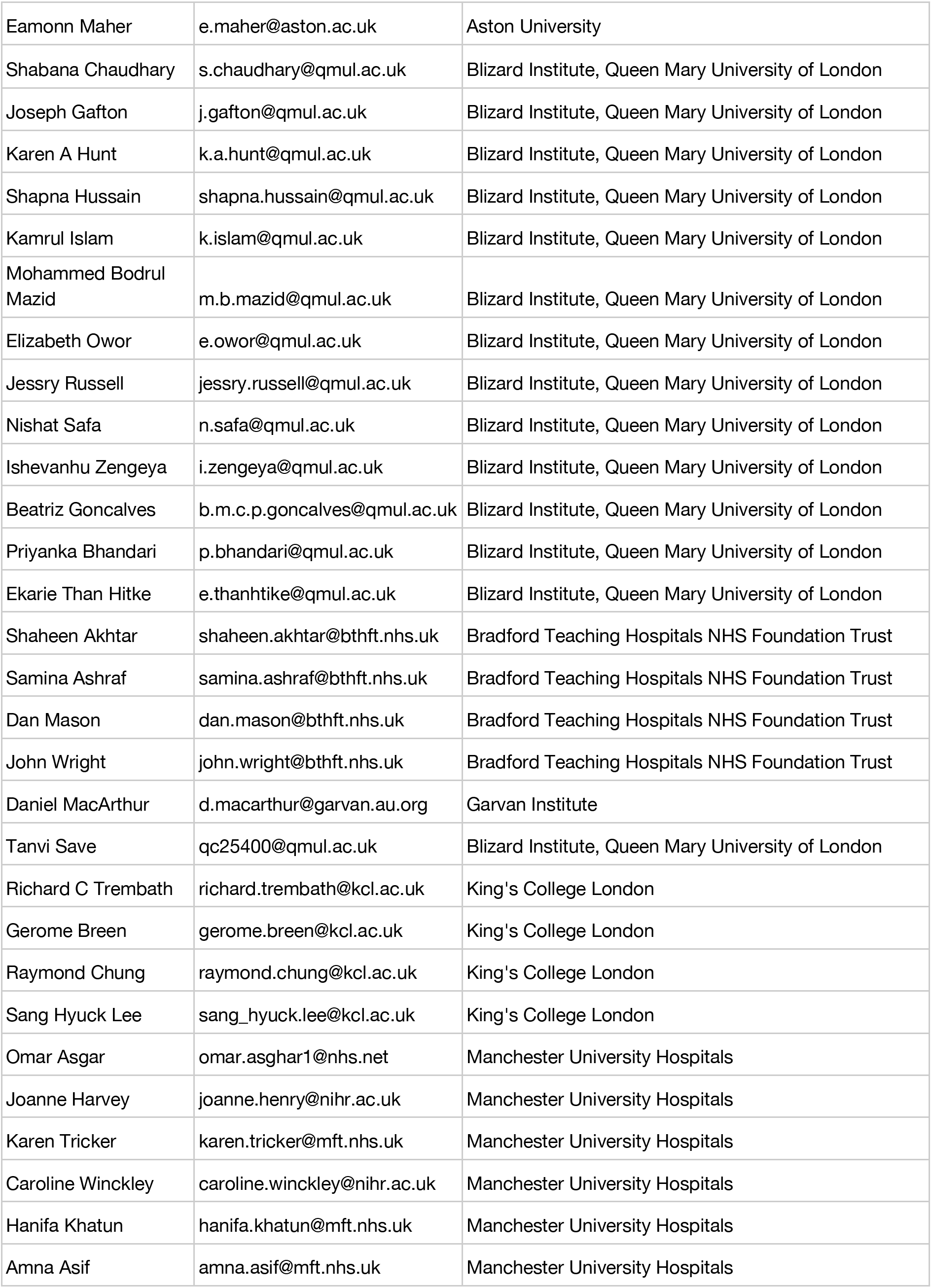
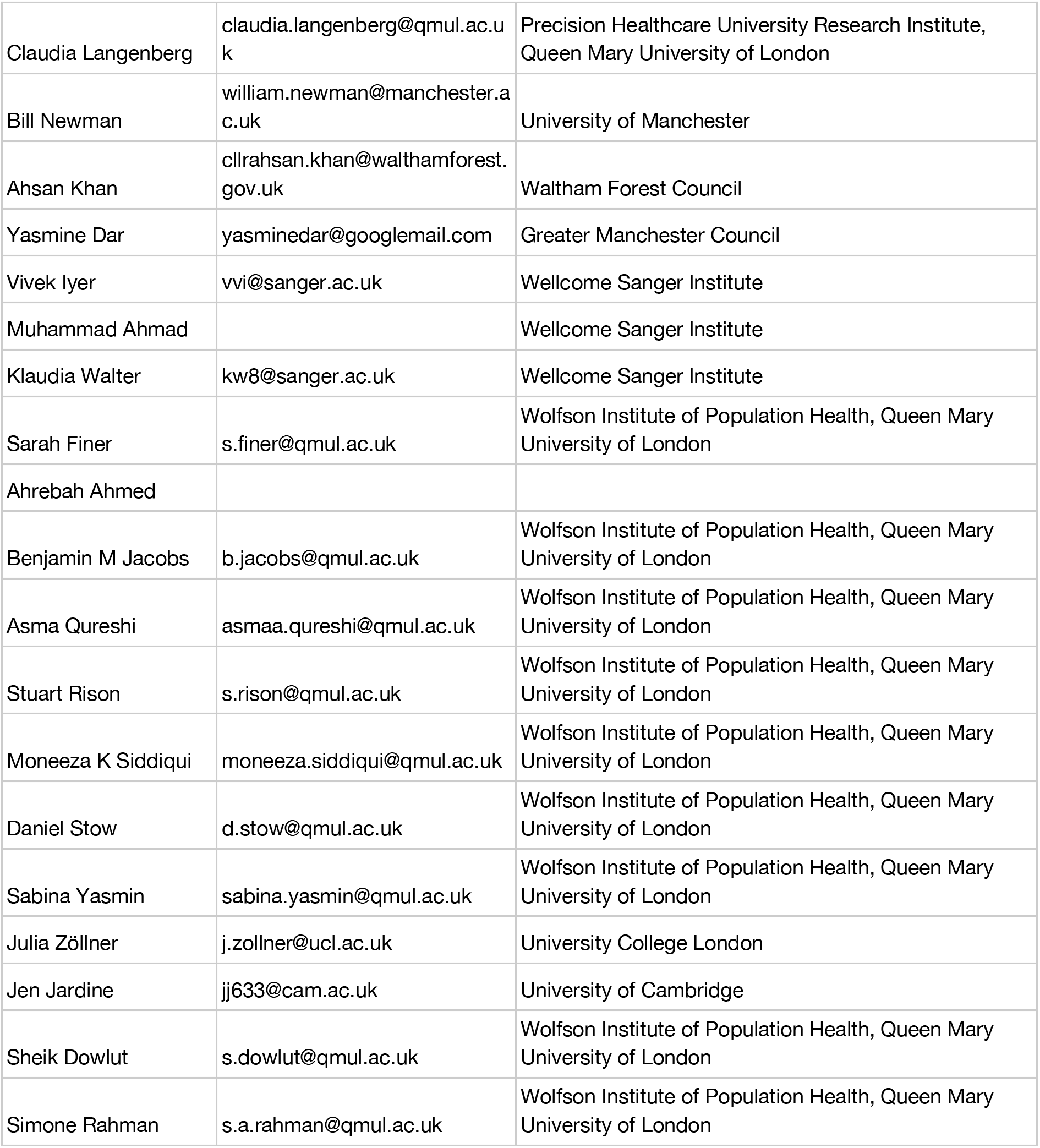

## Data availability

The genotype and phenotype for the UKB cohort described in this study were obtained from the UK Biobank resource under approved application number 69938. These are available, together with raw and processed single-cell data, to bona fide researchers upon approved application to the UK Biobank Access Management System.

The health and genetic data for the G&H cohort used in this study were obtained from the Genes & Health resource. These are available, together with raw and processed single-cell data, to registered researchers via the secure Genes & Health Trusted Research Environment following approval by the cohort’s data access committee.

The CellTypist cell type prediction models for celltype 2 and celltype 3 labels are available in the CARDINAL data central repository (data will be released upon publication) and through the ArcheTypist package for celltype 3.

## Code availability

The cell2clone package is available on GitHub (https://github.com/TeichLab/cell2clone) and ReadTheDocs (https://cell2clone.readthedocs.io). The ArcheTypist package is available on GitHub (https://github.com/TeichLab/ArcheTypist) and ReadTheDocs (https://archetypist.readthedocs.io). Code used for specific analyses described in this paper is available on GitHub (https://github.com/Teichlab/cardinal_blood_archetypes).

