## Extended Data Figures for "Multimodal population-scale cell atlases define archetypes and extremes of human blood"

**Extended Data Figure 1.** Associations between blood archetypes, demographics and clinical variables.

**Extended Data Figure 2.** Associations between blood archetypes and circulating haematopoietic progenitors.

**Extended Data Figure 3.** Regulons associated with cell composition trajectories.

**Extended Data Figure 4.** Benchmarking cell2clone in datasets with paired single-cell transcriptomic and VDJ sequencing data.

**Extended Data Figure 4**. Benchmarking cell2clone in datasets with paired single-cell transcriptomic and VDJ sequencing data.

**Extended Data Figure 5.** Clonality subgroups and donor-specific cell states associated with clonal expansions.

**Extended Data Figure 6.** Impact of mosaic chromosomal alterations and CHIP on cell states.

**Extended Data Figure 7.** Cell-state-resolved mosaic chromosomal alterations and subclonal evolution in CD5+ and exhausted B cells.

**Extended Data Figure 8.** Germline regulatory variation in CD5+ B cells.

**Extended Data Figure 9.** Classifying blood abnormalities and public PBMC datasets using the archetype model.

**Extended Data Figure 10**. ArcheTypist model to classify tissue composition subtypes.

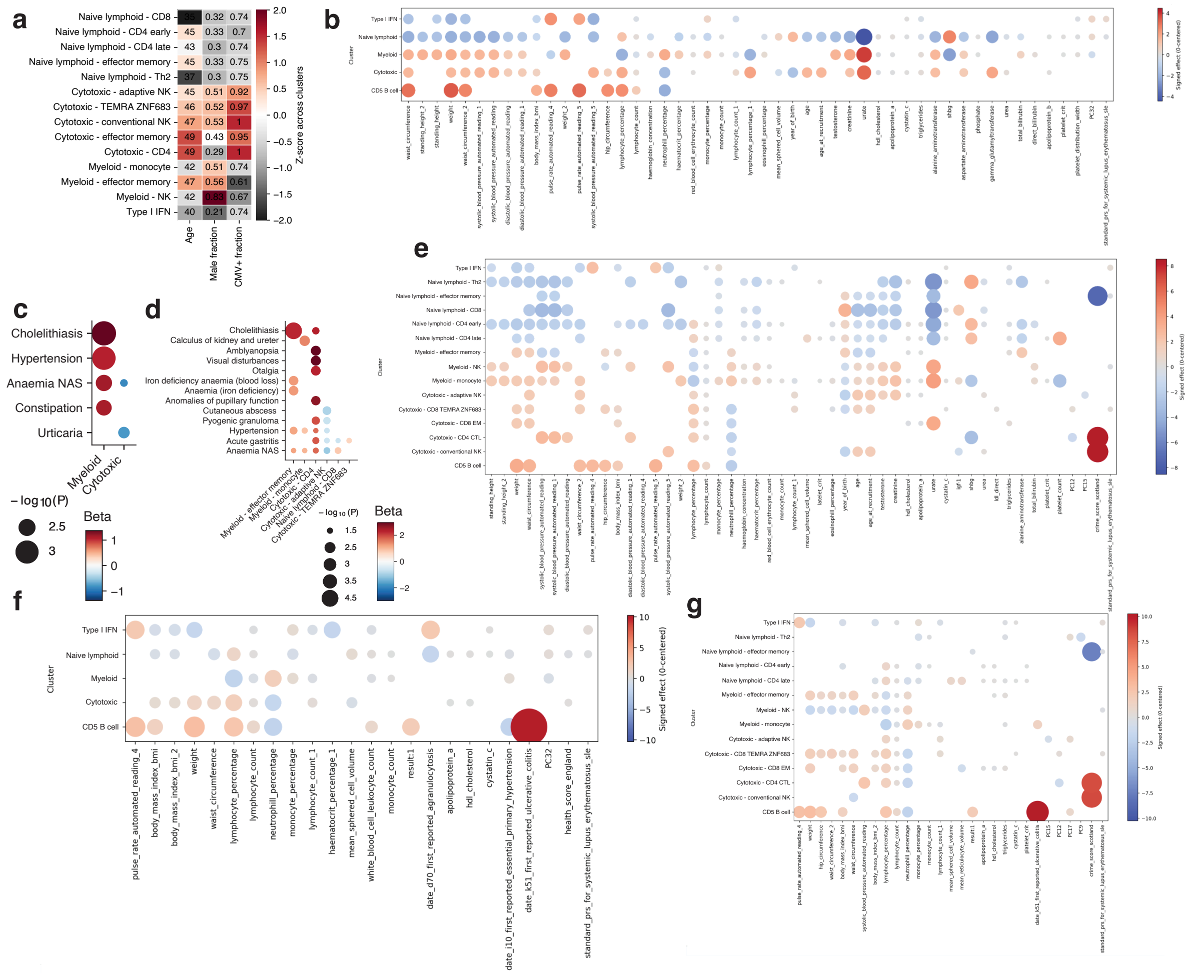


### Extended Data Figure 1. Associations between blood archetypes, demographics and clinical variables.

**a,** Median age, fraction of male sex and fraction of predicted CMV seropositive donors in blood composition subtypes in the G&H cohort shown as heatmap. **b,** Associations of blood archetypes with clinical and blood count variables from the UKB cohort without adjustment. Dot size indicates significance (-log10 P value) and colour indicates signed effect from the linear model. Only significant associations are shown. **c,** Associations of blood archetypes with clinical and blood count variables from the G&H cohort after adjustment for age and sex. Dot size indicates significance (-log10 P value) and colour indicates signed effect size. Only significant associations are shown. **d,** As in c but for blood composition subtypes. **e,** As in b but for blood composition subtypes. **f,** Associations of blood archetypes with clinical and blood count variables from the UKB cohort after adjustment for age, sex and ancestry. Dot size indicates significance (-log10 P value) and colour indicates signed effect from the linear model. Only significant associations are shown. **g,** As in f but for blood composition subtypes.

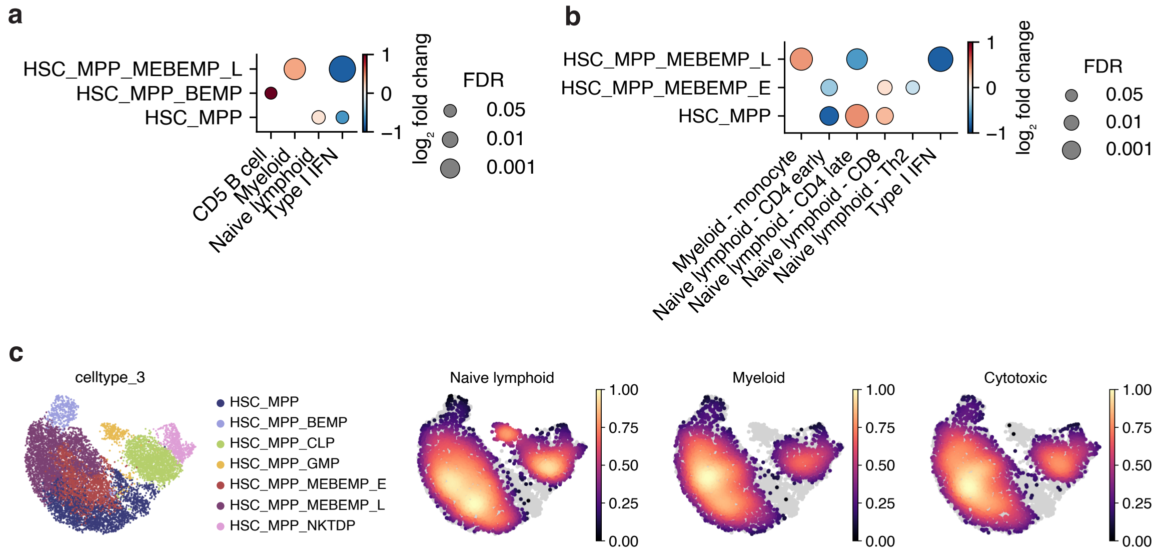


**Extended Data Figure 2. Associations between blood archetypes and circulating haematopoietic progenitors.**

**a,** Enrichment of circulating haematopoietic progenitor cell types in blood archetypes. Dot size indicates false discovery rate and colour indicates log2 fold change relative to all other archetypes. Only significant associations are shown (FDR<0.1 and P<0.05) **b,** Enrichment of circulating haematopoietic progenitor cell types in blood composition subtypes. Dot size indicates false discovery rate and colour indicates log2 fold change relative to all other subtypes. Only significant associations are shown (FDR<0.05). **c,** UMAP representations of circulating haematopoietic progenitor cells across the CARDINAL dataset, with cell type and density of three major archetypes coloured on the plots. Density plots illustrate enrichment of naive lymphoid archetype in earlier HSC_MPP cells, whereas myeloid archetype is enriched in later HSC_MPP_MEBEMP_L cells.

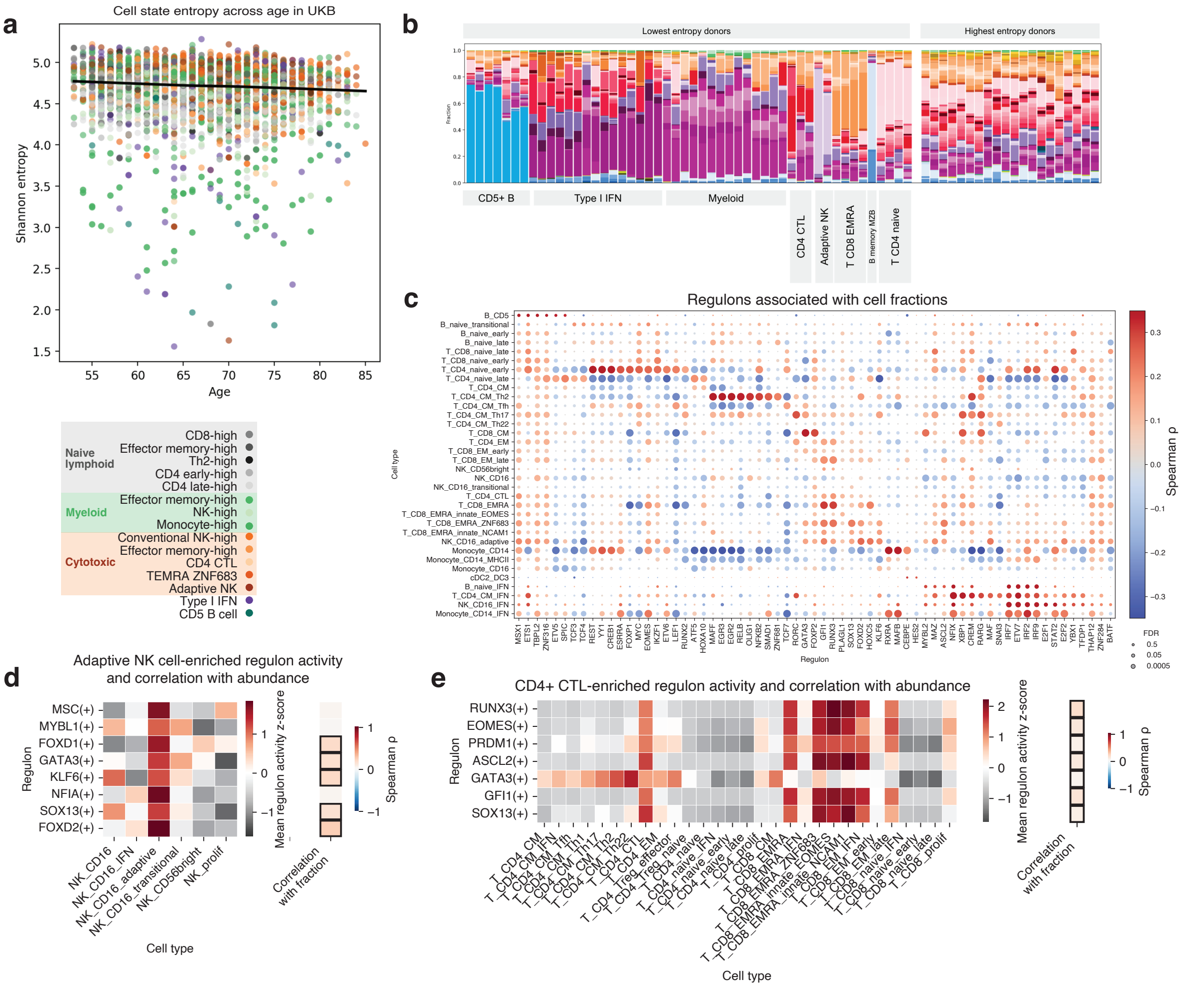


**Extended Data Figure 3. Cell state entropy and regulons associated with cell composition trajectories.**

**a,** Scatter plot comparing cell state entropy with age in the UKB cohort. Dot colour indicates blood composition subtype of each sample. **b,** Stacked barplot of cell type proportions (celltype_3) in donors with lowest or highest cell state entropy across the UKB cohort. The samples are hierarchically clustered based on cell type proportions and groups of samples with similar cell composition labelled by the archetype or characteristic cell type. **c,** Dot plot showing correlation of cell type fractions with regulon activity across all samples with at least 20 cells in the tested cell type. Dot colour indicates Spearman's correlation coefficient, with red colour indicating higher regulon activity associated with higher cell fraction out of all cells. Dot size indicates significance of the correlation shown as -log10(FDR). **d,** Regulons enriched in adaptive NK cells compared to other NK cell subsets. Heatmap shows the mean regulon activity z-score across samples in all cell types. Column on the right indicates correlation of regulon activity with cell fraction across samples, with colour denoting Spearman's correlation coefficient and significant correlations (FDR<0.05) indicated with a black border. **e,** As in d but for regulons enriched in CD4+ CTLs compared to other CD4+ and CD8+ T cell subsets.

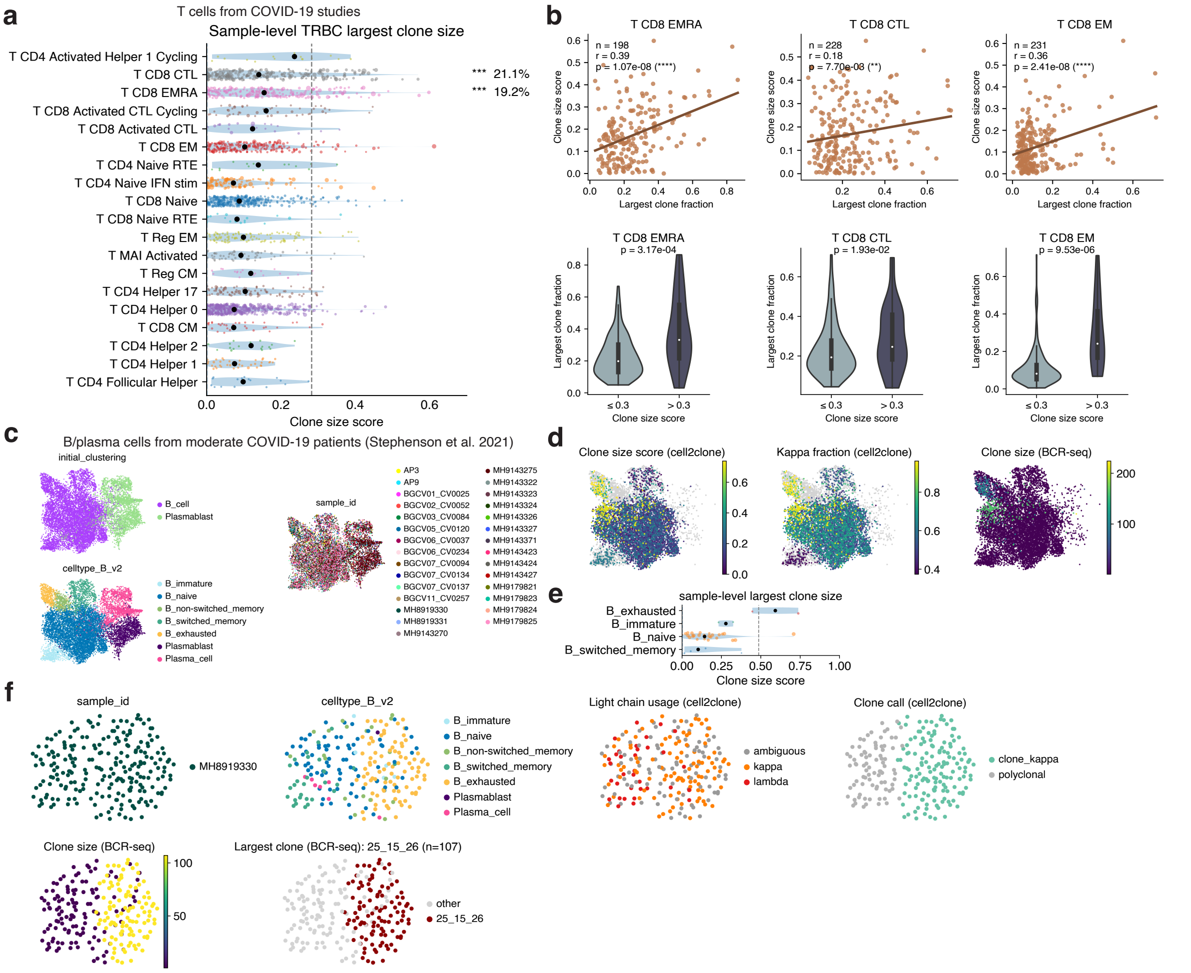


**Extended Data Figure 4. Benchmarking cell2clone in datasets with paired single-cell transcriptomic and VDJ sequencing data.**

**a,** Inferred cell2clone-derived dominant clone size across samples in T cell subtypes from COVID-19 datasets from the pan-infection atlas (Dratva et al. tcellatlas.org). Dots indicate individual samples, dot size corresponds to the fraction of the cell type out of the compartment in that sample, and black dots indicate median across samples. Only donors with at least 40 cells of each cell type are included in the analysis. Asterisks indicate significance based on the permutation test assessing presence of high-clonality samples in the cell type compared to non-clonal naive cells: *** FDR<0.001, ** FDR<0.01, * FDR<0.05. Percentages indicate samples above the threshold of clone size score defined based on non-clonal naive cells (97.5th percentile), indicated with a dashed line. **b,** Scatter plots and violin plots comparing cell2clone-derived clone size score against VDJ-seq-derived largest clone fraction per cell type in T cell subtypes from COVID-19 datasets from the pan-infection atlas. Scatter plots indicate number of samples, Pearson correlation coefficient and p value for correlation across samples. Violin plots show VDJ-seq-derived largest clone fraction stratified by cell2clone-derived clone size score using a cutoff (0.3) defined based on 97.5th percentile of the clone size score (0.28) in naive CD4+ T cells. P values are from a Mann-Whitney U test. Shown cell types are selected based on a significant proportion of clonal samples inferred by cell2clone (T CD8 EMRA, T CD8 CTL) and presence of individual samples with high clone size score (T CD8 EM) based on panel a. **c,** UMAP representations of B and plasma cells from moderate-severity COVID-19 patients from Stephenson et al., with coarse- and fine-grained cell type annotations and sample IDs coloured on the plots. Annotations and UMAP coordinates are from Stephenson et al. **d,** Cell2clone-derived inferred clone size score and kappa light chain fraction as well as BCR-seq-derived clone size coloured on single B cells. For clone size score and kappa light chain fraction, all B cells from the same sample and cell type have the same value. Plots demonstrate utility of cell2clone to correctly highlight regions of a cell embedding harbouring clonal expansions. **e,** Inferred cell2clone-derived dominant clone size across samples in B cell subtypes from the moderate-severity COVID-19 patients from Stephenson et al. Dots indicate individual samples, dot size corresponds to the fraction of the cell type out of the compartment in that sample, and black dots indicate median across samples. Only donors with at least 40 cells of each cell type are included in the analysis. Dashed line indicates the threshold of clone size score defined based on non-clonal naive cells (97.5th percentile). **f,** UMAP representations of B cells from a single sample from a moderate-severity COVID-19 patient from Stephenson et al., harbouring a large cell2clone-inferred clonal expansion in exhausted B cells. Coloured on the plots are sample ID, cell type, cell2clone-derived light chain usage, cell2clone-derived clone call status (based on enrichment of cells of the same light chain class in a Leiden cluster), BCR-seq-derived clone size, and BCR-seq-derived largest clone identity. Plots demonstrate utility of cell2clone to correctly identify clonal expansions at single-cell resolution.

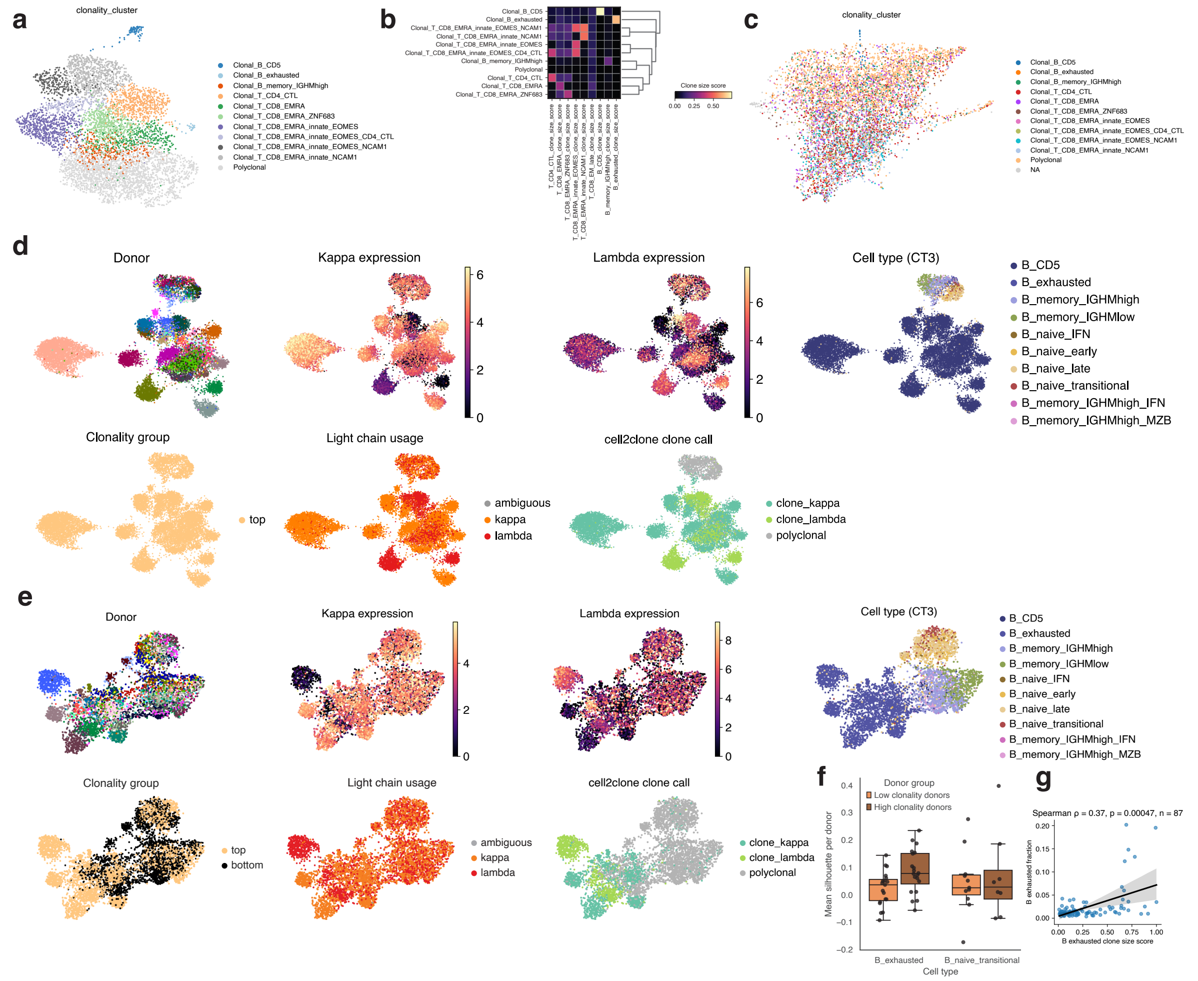


**Extended Data Figure 5. Clonality subgroups and donor-specific cell states associated with clonal expansions.**

**a,** UMAP and Leiden clustering-derived subtypes based on clone size scores inferred using cell2clone. Donors clustering together exhibit clonal expansions in the same cell types. **b,** Heatmap of mean clone size score of the indicated cell states across clonality-based subgroups. Subgroups are ordered using hierarchical clustering. **c,** Clonality-based subgroups coloured on a force-directed graph based on cell type composition. **d,** UMAP representations of B cells from donors with highest fractions of CD5+ B cells present out of all B cells. Coloured on the plots are donor identity, kappa (IGKC) and lambda (sum of IGLC genes) light chain gene expression, cell type, clonality group, light chain usage classification based on cell2clone and cell2clone clone call. Plots demonstrate donor-specific grouping of CD5+ B cells and a light chain usage pattern indicative of clonality, serving as the basis for cell2clone clonal inference. **e,** As in d but for atypical (age-associated, exhausted) B cells. **f,** Silhouette scores from cell2clone for atypical (age-associated, exhausted) B cells and transitional B cells from low and high-clonality (based on B_exhausted) donors based on the nearest neighbour space on the same data shown in e. Plot demonstrates higher silhouette score i.e. higher donor-specific grouping of transcriptomes of clonal atypical B cells than polyclonal atypical B cells. Transitional B cells are shown as control to indicate no difference as no difference in clonality is observed between the groups in this cell type. **g,** Scatter plot comparing fraction of exhausted B cells out of all cells in a sample and the cell2clone-derived clone size score in exhausted B cells. Only samples with at least 20 exhausted B cells are included in the analysis enable clonality estimates.

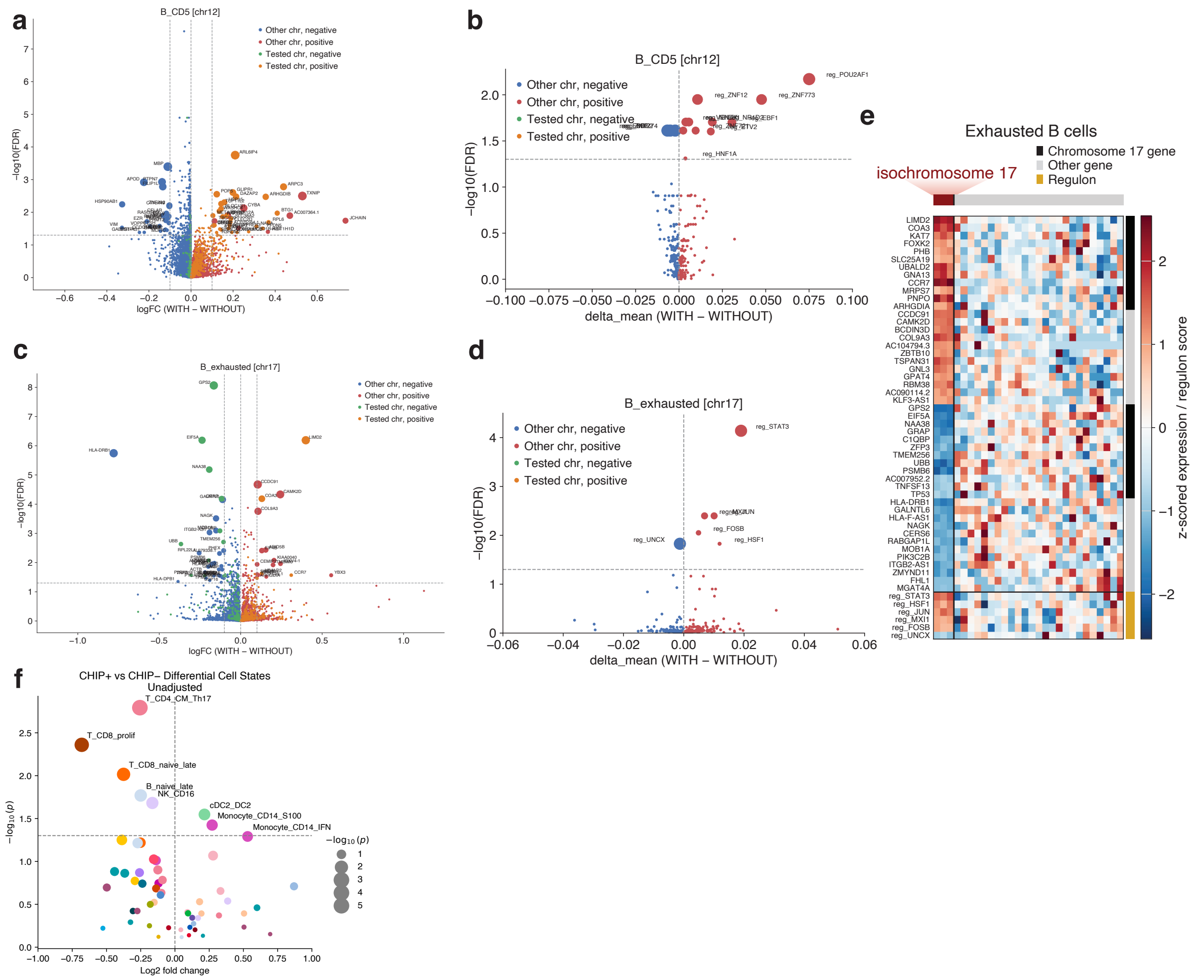


**Extended Data Figure 6. Impact of mosaic chromosomal alterations and CHIP on cell states.**

**a,** Volcano plot showing differentially expressed genes between CD5+ B cells from donors with or without a chromosome 12 amplification. **b,** Volcano plot showing differentially active regulons between CD5+ B cells from donors with or without a chromosome 12 amplification. **c,** Volcano plot showing differentially expressed genes between exhausted B cells from donors with or without an isochromosome 17 alteration. **d,** Volcano plot showing differentially active regulons between exhausted B cells from donors with or without an isochromosome 17 alteration. **e,** Heatmap showing significantly differentially expressed genes and differentially active regulons in exhausted B cells from donors with or without an isochromosome 17 alteration. **f,** Volcano plot showing differentially enriched cell types between donors harbouring CHIP and those without (no adjustment for age, sex and ancestry).

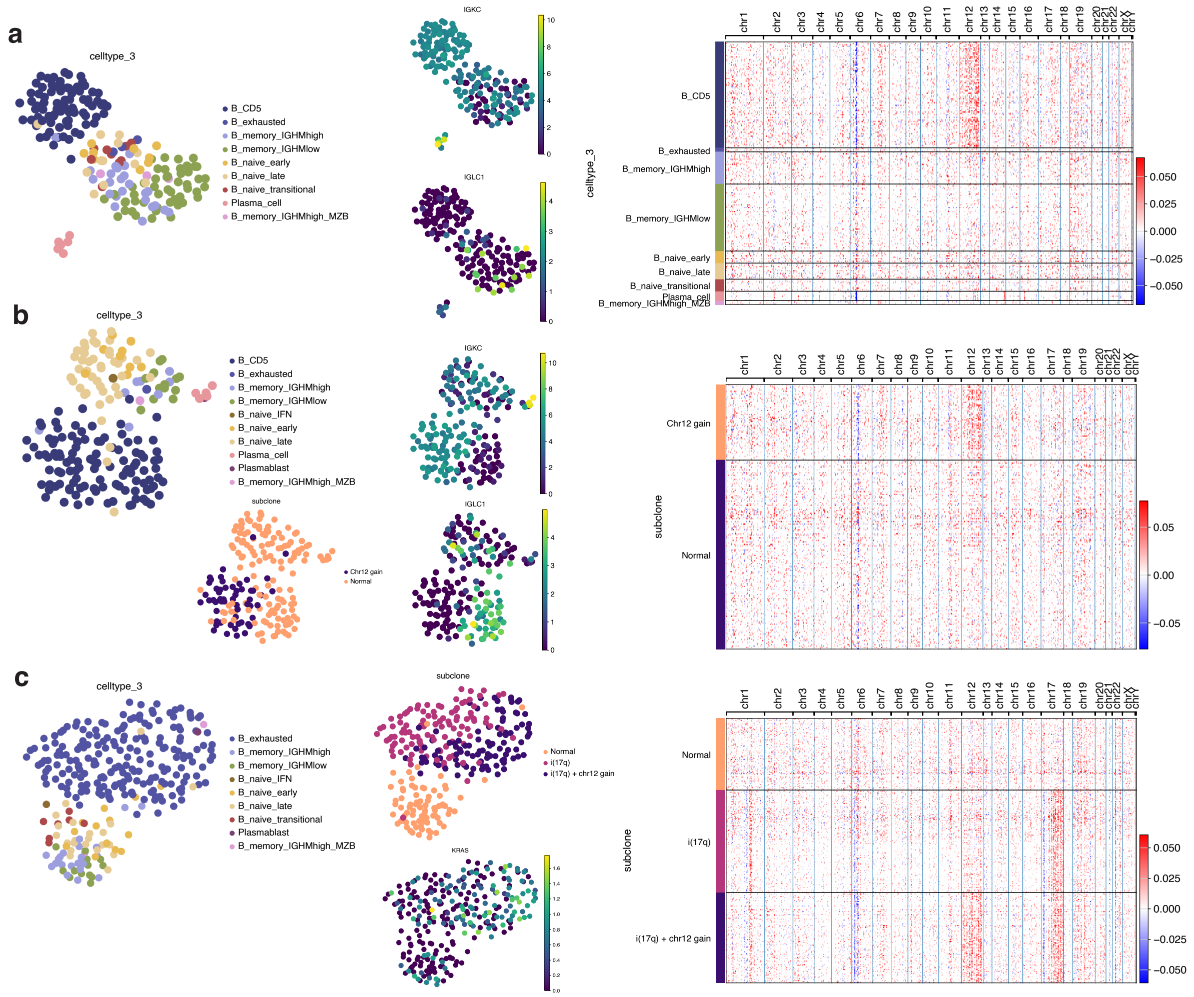


**Extended Data Figure 7. Cell-state-resolved mosaic chromosomal alterations and subclonal evolution in CD5+ and exhausted B cells.**

**a,** B and plasma cells from one sample harbouring inferred chromosome 12 trisomy in CD5+ cells. UMAPs show cell type annotation (celltype_3) and *IGKC* and *IGLC1* expression. Heatmap shows expression levels of genes ordered by chromosomes across all cells from the sample, grouped by celltype_3. **b,** B and plasma cells from one sample harbouring two inferred clones (kappa and lambda), with the kappa clone additionally showing chromosome 12 trisomy in CD5+ cells. UMAPs show cell type annotation (celltype_3), subclone status inferred based on CNA status and *IGKC* and *IGLC1* expression. Heatmap shows expression levels of genes ordered by chromosomes across all cells from the sample, grouped by CNA subclone status. **c,** B and plasma cells from one sample harbouring inferred isochromosome 17 and chromosome 12 trisomy in exhausted/atypical B cells. UMAPs show cell type annotation (celltype_3), subclone status inferred based on CNA status, and expression of KRAS, a gene differentially expressed in cells with inferred chromosome 12 trisomy.. Heatmap shows expression levels of genes ordered by chromosomes across all cells from the sample, grouped by CNA subclone status.

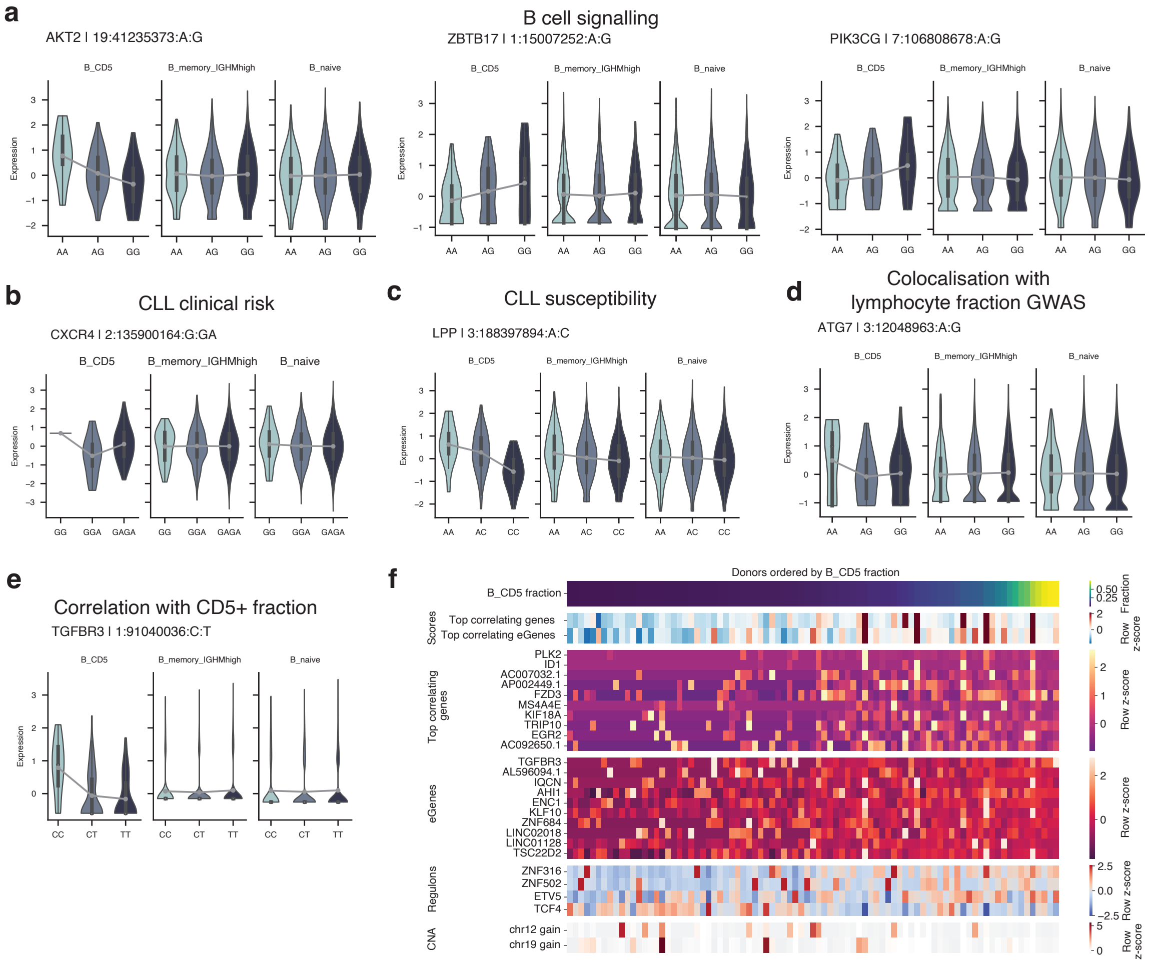


**Extended Data Figure 8. Germline regulatory variation in CD5+ B cells.**

**a,** Examples of dosage effects of CD5+ B cell-specific eQTLs involving genes mediating B cell receptor signalling. **b,** Example of dosage effect of a CD5+ B cell-specific eQTL involving *CXCR4,* a gene associated with CLL clinical risk. **c,** Example of dosage effect of a CD5+ B cell-specific eQTL involving *LPP*, a gene linked to CLL susceptibility in GWAS studies. **d,** Example of dosage effect of a CD5+ B cell-specific eQTL involving *ATG7*, colocalising with lymphocyte fraction in GWAS. **e,** Example of dosage effect of a CD5+ B cell-specific eQTL involving *TGFBR3*, whose expression correlates with the fraction of CD5+ B cells across donors. **f,** Heatmap of top genes, eGenes and regulons correlating with CD5+ B cell fraction in samples with at least 10 CD5+ B cells.

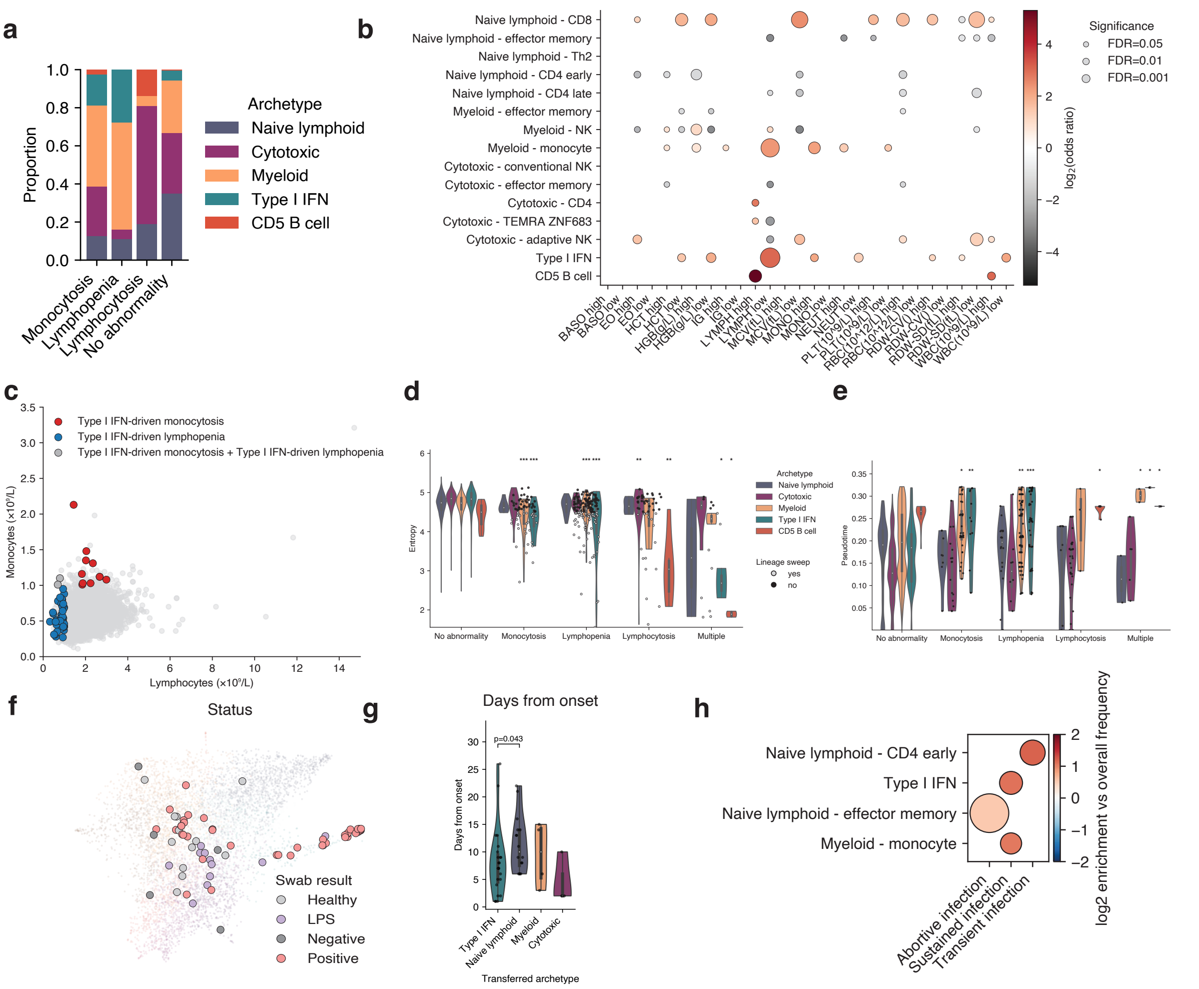


### Extended Data Figure 9. Classifying blood abnormalities and public PBMC datasets using the archetype model.

**a,** Distribution of blood archetypes in blood count abnormalities shown as a stacked bar plot indicating the proportion of donors with each blood archetype stratified by the blood count abnormality. **b,** Associations of blood composition subtypes across all blood count abnormalities, defined as low or high counts of various CBC parameters based on clinical reference ranges. Dot colour indicates log2 odds ratio of the association, and dot size indicates significance. Only significant associations are shown. **c,** Scatter plot comparing lymphocyte and monocyte count from CBC across the cohort, with type I IFN-related archetype model-derived abnormality subclasses coloured. Red colour indicates type I IFN-driven monocytosis, blue colour indicates type I IFN-driven lymphopenia, and dark grey dots indicate a combination of the two. Plot demonstrates the primarily mutually exclusive occurrence of type I IFN-driven monocytosis and lymphopenia. **d,** Cell state entropy stratified by archetype and blood abnormality status shown as a violin plot. Dots indicate individual samples (shown only for blood abnormalities), and dot colour indicates lineage sweep status. Asterisks indicate significance of entropy decrease in samples with a blood count abnormality compared to samples of the same archetype without abnormalities. **e,** Blood composition pseudotime stratified by archetype and blood abnormality status shown as a violin plot. Dots indicate individual samples (shown only for blood abnormalities), and dot colour indicates lineage sweep status. Asterisks indicate significance of pseudotime increase in samples with a blood count abnormality compared to samples of the same archetype without abnormalities. **f,** Association of archetypes with COVID-19 swab results of samples from the COVID-19 PBMC study (Stephenson et al. 2021). Swab result status is coloured on the blood composition map. **g,** Violin plot of the days from onset of samples from the COVID-19 PBMC study (Stephenson et al. 2021) stratified by archetypes. P values are shown for the significant differences between archetypes based on a Mann-Whitney U test. **h,** Enrichment of blood composition subtypes from any time point in infection outcome categories in the COVID-19 human challenge (Lindeboom et al. 2024).

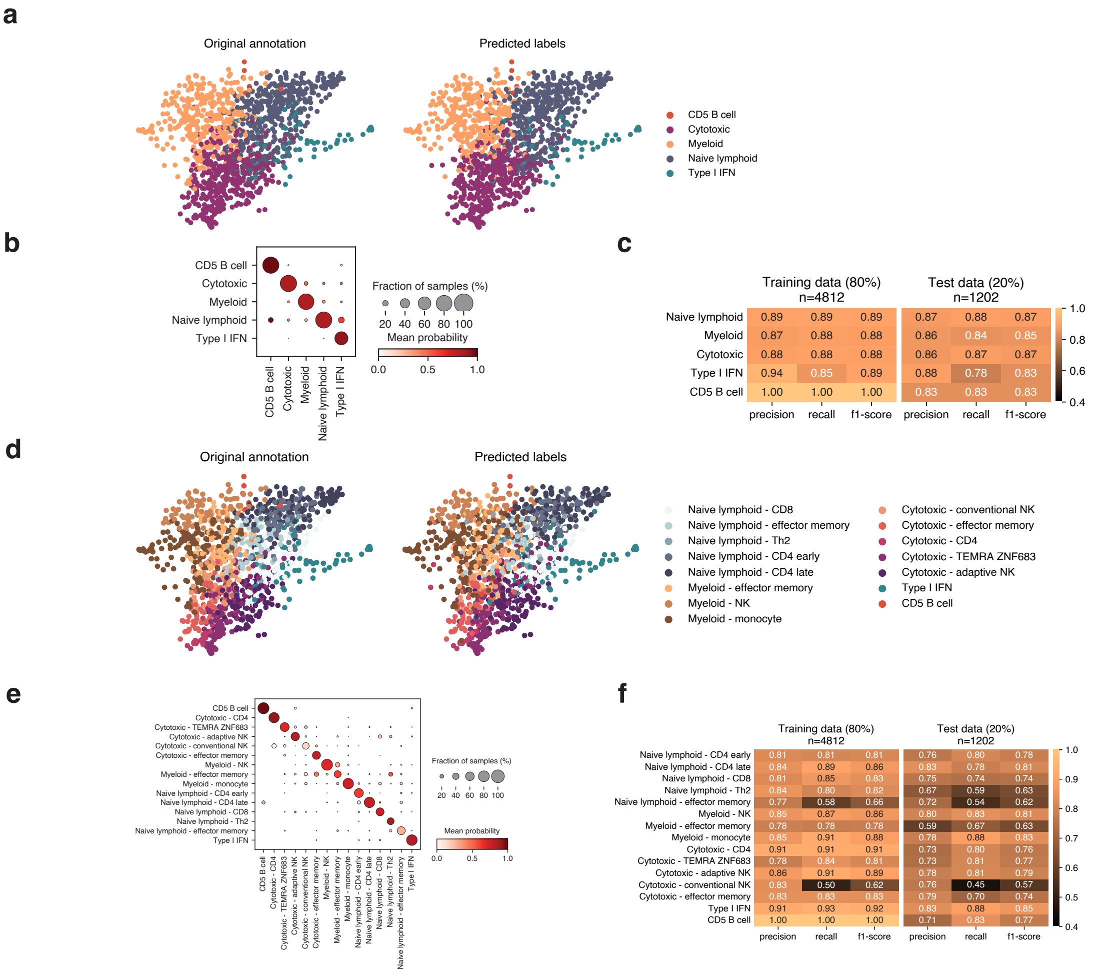


**Extended Data Figure 10. ArcheTypist model to classify blood archetypes and blood composition subtypes.**

**a,** Original and predicted archetype status coloured on the cell type composition-based force-directed graph in the held-out test set. **b,** Dot plot showing probability of the predicted archetypes grouped by original archetypes in the held-out test set. **c,** Heatmap showing precision, recall and f1-score for predicting each archetype in the training and test sets. **d,** Original and predicted cell composition subtype status coloured on the cell type composition-based force-directed graph in the held-out test set. **e,** Dot plot showing probability of the predicted cell composition subtypes grouped by original subtypes in the held-out test set. **f,** Heatmap showing precision, recall and f1-score for predicting each cell composition subtype in the training and test sets.
