## Supplementary Figures for "Multimodal population-scale cell atlases define archetypes and extremes of human blood"

**Supplementary Figure 1.** Cell type fractions coloured on the blood composition map.

**Supplementary Figure 2.** Cell state-enriched regulons across 79 blood cell states.

**Supplementary Figure 3.** Gene regulatory networks underlying blood cell states split by lineage.

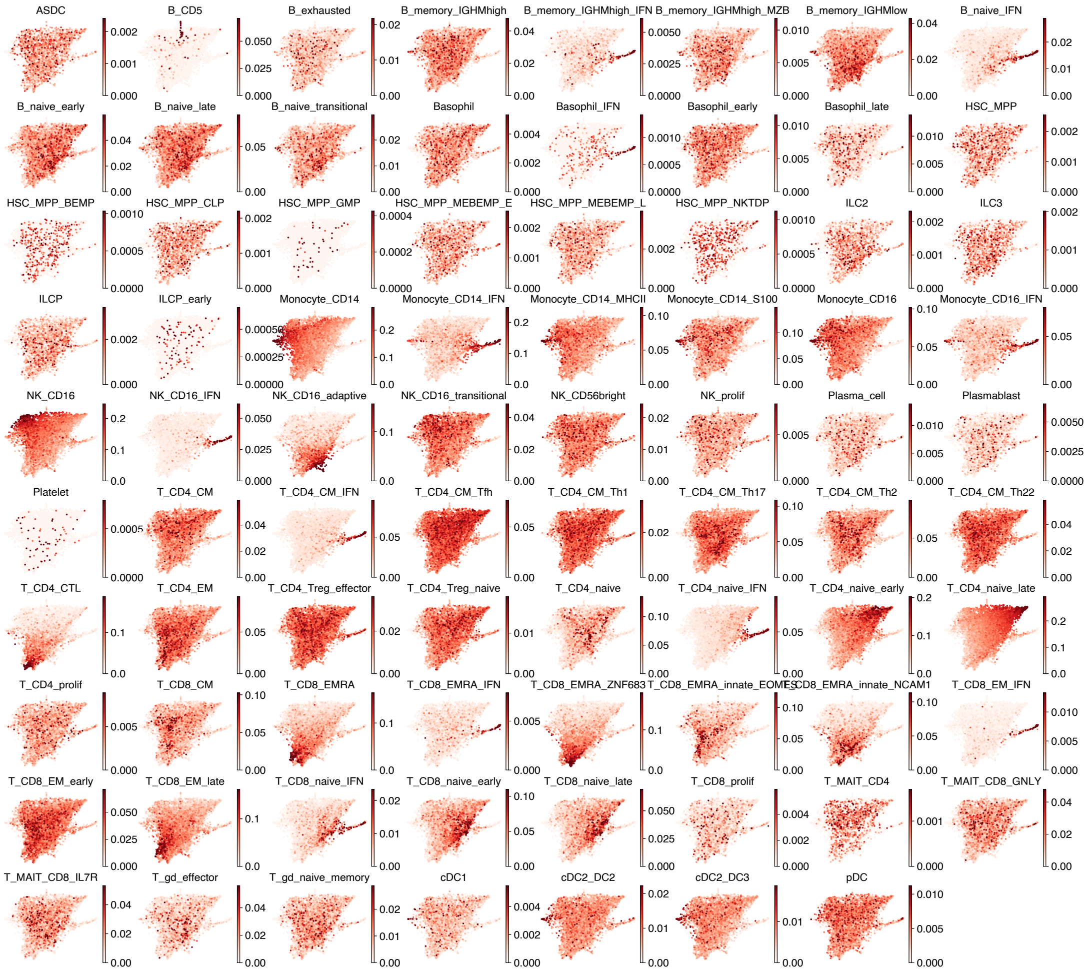


**Supplementary Figure 1. Cell type fractions coloured on the blood composition map.**

Fractions of each CT3 cell type out of all cells in a sample coloured on a force-directed layout based on cell state fractions at the CT3 level across 6,014 samples in the UKB and G&H cohorts. Each dot indicates a blood sample from one donor.

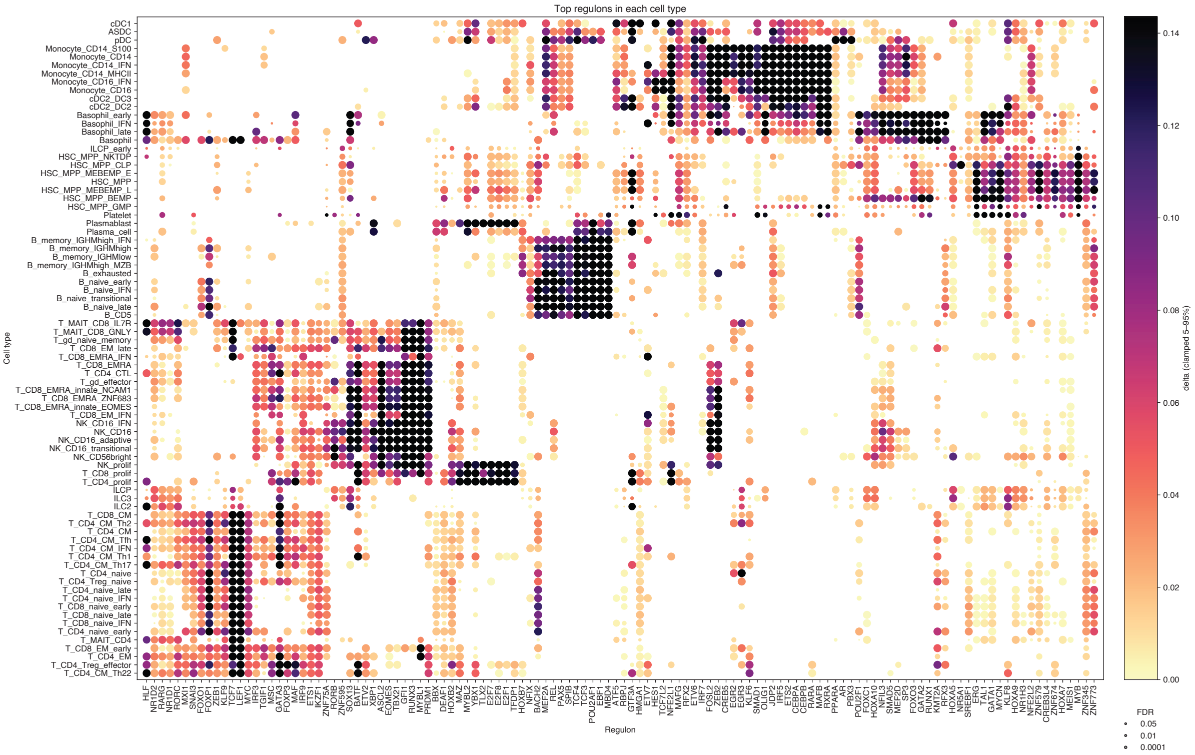


**Supplementary Figure 2. Cell state-enriched regulons across 79 blood cell states.**

Dot plot of differentially active regulons between cell types at the CT3 level across all cell types. Dot size indicates significance of the differential activity between sample-level pseudobulks of each cell type compared to all other cell types, shown as -log10(FDR), and colour indicates the difference in mean regulon scores between the given cell type and all other cell types. Regulons and cell types are hierarchically clustered.

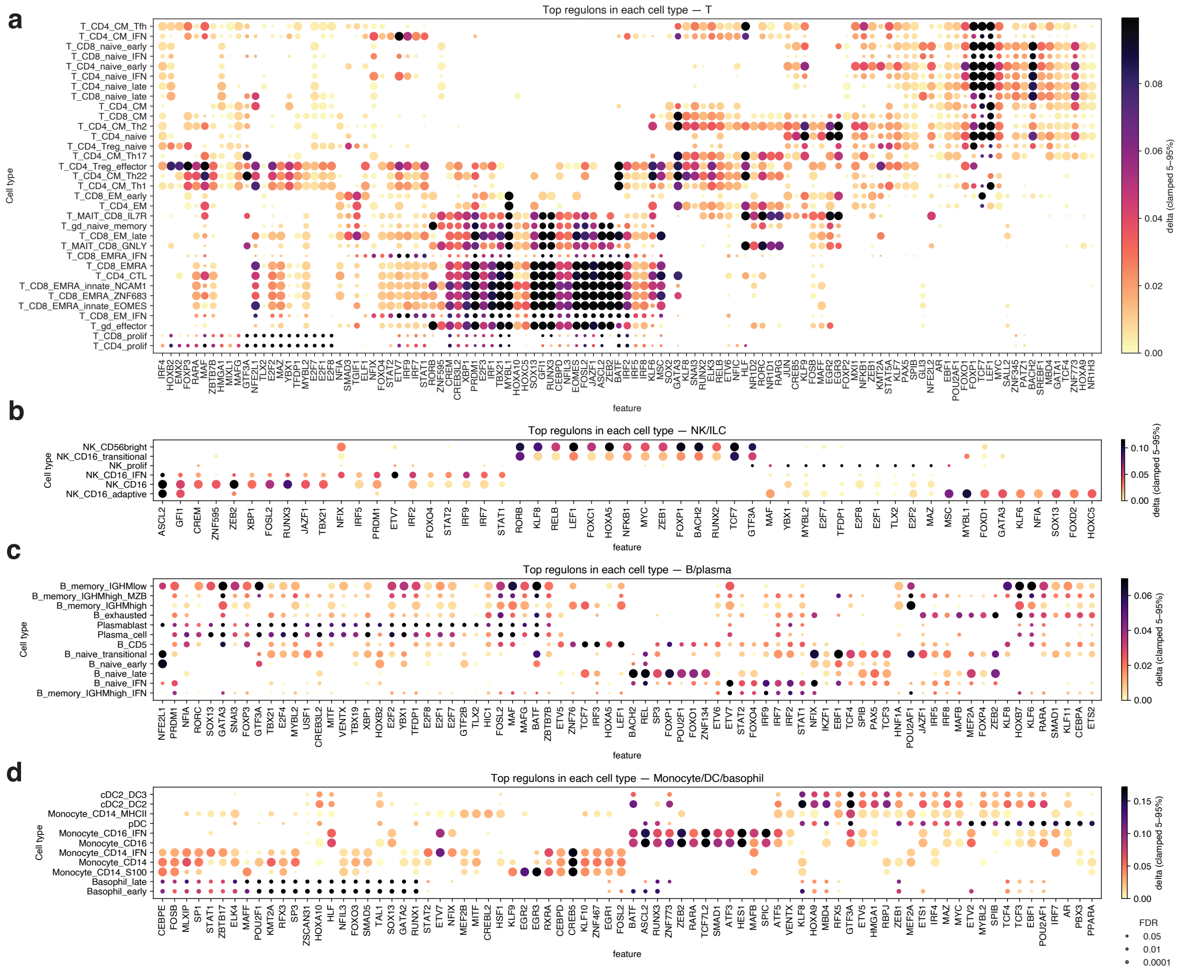


**Supplementary Figure 3. Gene regulatory networks underlying blood cell states split by lineage.**

**a,** Dot plot of differentially active regulons between cell types at the CT3 level for T cells. Dot size indicates significance of the differential activity between sample-level pseudobulks of each cell type compared to all other cell types, shown as -log10(FDR), and colour indicates the difference in mean regulon scores between the given cell type and all other cell types. Regulons and cell types are hierarchically clustered. **b,** As in a for NK/ILC. **c,** As in a for B/plasma. **d,** As in a for Monocyte/DC/basophil.
